# ERBB4 deficiency promotes atrial myopathy underlying the atrial fibrillation substrate

**DOI:** 10.64898/2026.05.26.26354173

**Authors:** Naoko Yamaguchi, John Santucci, Seok Jae Hong, Alexander Ferrena, Florencia Schlamp, David K. Willett, Charlotte J. Casdin, Peter S. Park, Xianming Lin, Junhua Xiao, Samantha Hall, John Barnard, Jonathan S. Achter, Konstantin Kahnert, Alicia Lundby, Mina K. Chung, David R. Van Wagoner, David S. Park

## Abstract

**Background:** Atrial fibrillation (AF) is a leading cause of stroke, cardiovascular morbidity, and mortality. Atrial myopathy, characterized by progressive metabolic, electrical, and structural changes, creates the arrhythmogenic substrate that drives AF. Defining the key drivers of atrial myopathic processes is essential for targeted therapies that can mitigate AF progression. Here we explore how reduced ERBB4 expression contributes to the development of left atrial myopathy.

**Methods:** We analyzed the Cleveland Clinic Biobank to compare left atrial *ERBB4* levels in patients grouped by AF diagnosis. To investigate the impact of reduced ERBB4 levels on atrial tissue substrate, we created mouse models of cardiac-specific *Erbb4* deficiency using Mlc2a (myosin light chain 2a)-Cre. Comprehensive physiological assessments were performed. Transcriptomic analyses of the left atrium were performed in an *Erbb4* haploinsufficient mouse model and compared with human atrial datasets. Molecular validation of key dysregulated pathways was performed.

**Results:** We found that left atrial *ERBB4* levels are reduced in patients with AF. Adult cardiomyocyte-specific *Erbb4* heterozygous (*Erbb4*^fl/+^;Mlc2a-Cre) mice exhibited prolonged P-wave duration in the absence of ventricular dysfunction. Left atrial transcriptomic analysis in *Erbb4* haploinsufficient mice showed upregulation of pathways related to fibrosis, apoptosis, and coagulation, and downregulation of pathways related to fatty acid metabolism and mitochondrial function, mirroring changes observed in pressure overload mouse models. A cross-species transcriptomic comparison revealed significant overlap between *ERBB4*-correlated gene expression and functional pathways in adult human atria and mice with *Erbb4* haploinsufficiency. Validating the transcriptomic data, protein and functional assays demonstrated increased fibrosis, apoptosis, and oxidative stress in the mutant left atrial tissue.

**Conclusion:** Left atrial *ERBB4* levels are reduced in AF patients. A mouse model of *Erbb4* deficiency and human atrial transcriptomic analyses highlight a role for ERBB4 in supporting normal atrial metabolism while protecting against inflammation, apoptosis, and fibrosis.

## Introduction

Large-scale genome-wide association studies (GWAS) have been instrumental in identifying gene variants that confer increased risk of atrial fibrillation (AF).^1,2^ Integrating these genetic insights with established disease networks can clarify the mechanisms by which electrical, structural, metabolic, and neurohormonal changes transform normal atrial tissue into a disordered substrate primed for arrhythmia. Initially, AF often presents as self-terminating, paroxysmal episodes, but with progressive atrial remodeling, AF typically transitions to persistent forms.^3,4^ Despite advances in AF genetics, the mechanisms by which individual genes drive atrial remodeling and subsequent AF development and progression remain poorly defined. To date, three variants located within an intron of the erb-b2 receptor tyrosine kinase 4 (*ERBB4*) gene on human chromosome 2q34 have been associated with AF.^1,5–7^ ERBB4 has been implicated in myocardial cell differentiation,^8,9^ trabeculation during ventricular chamber formation,^8,10^ and cardiomyocyte proliferation in the mature heart.^11^ While *ERBB4* signaling has a fundamental role in normal cardiac development, its contribution to the pathogenesis of AF in the adult heart has not been defined.

ERBB4 functions as a transmembrane receptor tyrosine kinase that heterodimerizes with the non-receptor tyrosine kinase ERBB2.^12^ ERBB4 mediates neuregulin1 (NRG1) signaling from endocardial cells to cardiomyocytes.^13,14^ In animal studies, constitutive *Nrg1*, *Erbb2* and *Erbb4* deficient mice exhibit embryonic lethality by embryonic day 10.5–11 due to severe cardiac defects with prominent failure of myocardial trabeculation.^8,15,16^ Conditional *Erbb4* and *Erbb2* deficient mice generated using ventricular myosin light chain Cre-recombinase (MLC2v-Cre) exhibited severely dilated ventricular chambers and abnormal conduction, supporting the hypothesis that ERBB4 signaling in cardiomyocytes is specifically required for normal chamber development.^17,18^ With respect to atrial physiology, homozygous *l11Jus8* mutant mice, which carry a hypomorphic missense mutation of *Erbb2* generated by random chemical mutagenesis, exhibited distended atria and severe atrial conduction defects, ultimately resulting in midgestational lethality between embryonic days 12.5 and 13.^19^ While NRG1/ERBB4-ERBB2 signaling plays an important role in the development of the cardiac chambers, the transcriptional networks and functional pathways regulated by ERBB4 in the atrium remain incompletely defined, and its role in atrial remodeling is unknown.

We investigated the relationship between AF and *ERBB4* expression in human left atrial samples using the Cleveland Clinic Biobank.^20^ We then generated a cardiomyocyte-specific *Erbb4* deficiency mouse model using atrial myosin light chain Cre-recombinase (MLC2a-Cre). Integrated molecular, transcriptional, and functional assessments of *Erbb4* heterozygous (*Erbb4*^fl/+^Mlc2a^Cre^) mice reveal how reduced ERBB4 expression promotes pathologic atrial remodeling through its impact on metabolism, inflammation, coagulation, and fibrotic pathways.

## Methods

Detailed materials and methods are described in Supplemental Materials. All animal experiments were approved by the institutional animal care and use committee of New York University Grossman School of Medicine (PROTO201900150), and the animals received humane care in accordance with the US National Institutes of Health Guide for the Care and Use of Laboratory Animals. The human study protocol in this report was approved by the institutional review board of the Cleveland Clinic (2018-1501), and all patients provided informed consent for research use of discarded atrial tissue.

### Data Availability

The RNA-seq data from *Erbb4* mutant mouse atria are deposited in NCBI Gene Expression Omnibus (GEO) database (GSE288616). The RNA-seq data from human left atria in Cleveland Clinic Biobank are deposited in the GEO database (GSE69890).^20^ The RNA-seq data from cardiac pressure-overloaded mouse atria were obtained from GSE322182.^21^ The single nucleus RNA-seq data from adult human hearts were obtained from a previously published dataset (E-MTAB-12916).^22^ The RNA-seq data from human right atria were retrieved from Genotype-Tissue Expression project (GTEx) Portal.^23^

### Statistical Analysis

GraphPad Prism v10.4.0 (GraphPad Software, Boston, MA) was used for statistical comparisons except for sequencing data. Quantitative data were first evaluated by the Shapiro-Wilk test for normal distribution. Where appropriate, Student’s t-test was used for comparisons between two groups. One-way analysis of variance (ANOVA) with Tukey’s post-hoc test was performed for comparisons involving more than two groups. The *P* value < 0.05 was considered statistically significant. Values are presented as mean ± SD unless otherwise stated in the figure legend. Additional details specific to the analysis of RNA-seq data are described in the Supplemental Material.

## Results

### ERBB4 is downregulated in patients with persistent AF

To define cell type-specific *ERBB4* expression in the adult human heart, we analyzed a publicly available cardiac single-cell RNA-seq dataset.^22^ *ERBB4* is most highly expressed in atrial cardiomyocytes compared to other cell types found in the heart (Supplemental Figure S1A). This observation is consistent with previous studies of mouse embryonic hearts.^8^ Furthermore, consistent with human single-cell RNA-seq findings, ERBB4 protein levels in the adult mouse heart are higher in the atria than the ventricles (Supplemental Figure S1B).

We next quantified *ERBB4* expression levels in human left atrial samples from patients with and without AF utilizing the Cleveland Clinic Biobank, a repository of samples from 251 unique patients who underwent cardiac surgery.^20,21^ RNA-seq revealed that *ERBB4* levels are reduced in patients with AF compared to those in sinus rhythm at the time of surgery (*P*<0.0001, Figure 1A). We also investigated *ERBB4* levels by clinical AF subtype based on chart review of cardiac rhythm documentation. Patients with persistent AF had significantly lower *ERBB4* expression in the left atrium (LA) compared to those with no documented history of AF or paroxysmal AF (*P*=0.0252, Figure 1B).

**Figure 1.**
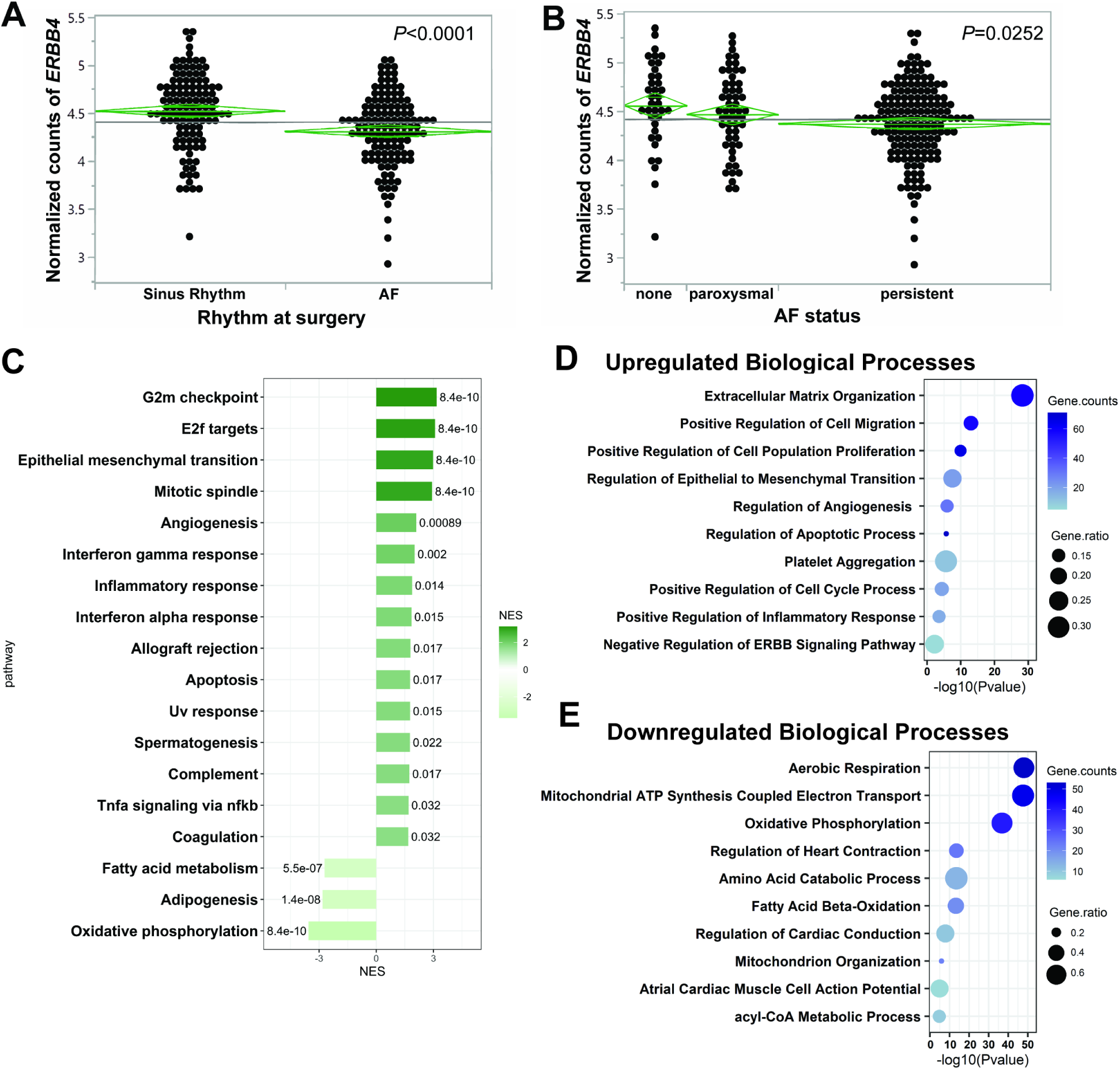
Left atrial *ERBB4* expression is reduced in patients with AF and pressure overloaded mouse models. **A**, *ERBB4* expression in left atrial appendage from RNAseq dataset in patients with sinus rhythm versus atrial fibrillation (AF) at time of surgery (n = 244). Green diamond represents group mean and 95% confidence intervals. Groups were compared by pooled *t*-test. **B**, *ERBB4* expression in the left atrial appendage of patients with chart diagnosis of paroxysmal AF, persistent AF, or neither (n = 251). Green diamond represents group mean and 95% confidence intervals. Groups were compared by one-way analysis of variance. **C**, Bar plot of significantly enriched hallmark pathways identified by Gene Set Enrichment Analysis with 2165 commonly differentially expressed genes (DEGs) in the cardiac pressure-overload mouse models by transverse aortic constriction banding and angiotensin II infusion for 2 weeks. Number next to the bar represents adjusted *P* value per pathway. Previously published dataset (GSE322182) was reanalyzed. NES, normalized enrichment score. **D and E**, Dot plots of significantly enriched upregulated (D) and downregulated (E) GO biological processes identified from the common DEGs in the cardiac pressure-overload mouse models. Gene counts indicate the number of input DEGs annotated to each GO term. Gene ratio is the proportion of input DEGs relative to all annotated genes of each term.

Reflecting the expression patterns observed in patients with clinical AF, our previous work established that cardiac pressure overload induced by transaortic constriction banding (TAC) or angiotensin II (AngII) infusion significantly reduces ERBB4 levels in the murine LA.^21^ To better understand the functional pathways consistently altered in the diseased LA, we analyzed our previously published LA RNA-seq datasets from TAC-banded and AngII-treated mice. The common differentially expressed genes (DEGs) between the two models were subjected to gene set enrichment analysis and Gene Ontology (GO) analysis, which revealed that cardiac pressure overload suppressed pathways involved in energy metabolism and enhanced pathways related to inflammation, apoptosis, coagulation, and extracellular matrix (ECM) organization (Figure 1C–E).

These data demonstrate that ERBB4 expression is reduced in human AF and in mouse models of pressure-overloaded LA, and that left atrial tissue with low ERBB4 expression is characterized by downregulated metabolic pathways and upregulated inflammatory, apoptotic, and coagulation pathways. Given these observations, we hypothesized that optimal ERBB4 signaling in atrial myocytes is required to maintain normal atrial structure and function in the adult heart.

### ERBB4 deficiency leads to atrial conduction slowing

To assess the impact of ERBB4 deficiency on atrial physiology, we created a cardiomyocyte-specific *Erbb4* deficient mouse model that selectively targets *Erbb4* exon 2 for deletion using MLC2a-Cre (*Myl7^tm1(cre)Krc^*, hereafter referred to as Mlc2a^Cre^).^21,24^ Tissue-specific Cre recombinase genotyping confirmed that the genetic deletion of *Erbb4* exon 2 only occurred in the presence of Mlc2a^Cre^-mediated excision (Figure 2A). *Erbb4* knockout mice (*Erbb4*^fl/fl^Mlc2a^Cre^) die *in utero* at approximately embryonic day (E) 11 due to severe ventricular trabeculation defects (data not shown), consistent with a previous report of germline *Erbb4* null mice.^8^ *Erbb4* heterozygous mice (*Erbb4*^fl/+^Mlc2a^Cre^) are viable and grossly indistinguishable from littermate controls genotyped as *Erbb4*^fl/+^ and Mlc2a^Cre^ (hereafter referred to as controls). *Erbb4* expression in the LA was significantly reduced at the RNA (Figure 2B) and protein levels (Figure 2C, D) in adult *Erbb4*^fl/+^Mlc2a^Cre^ mice compared to controls.

**Figure 2.**
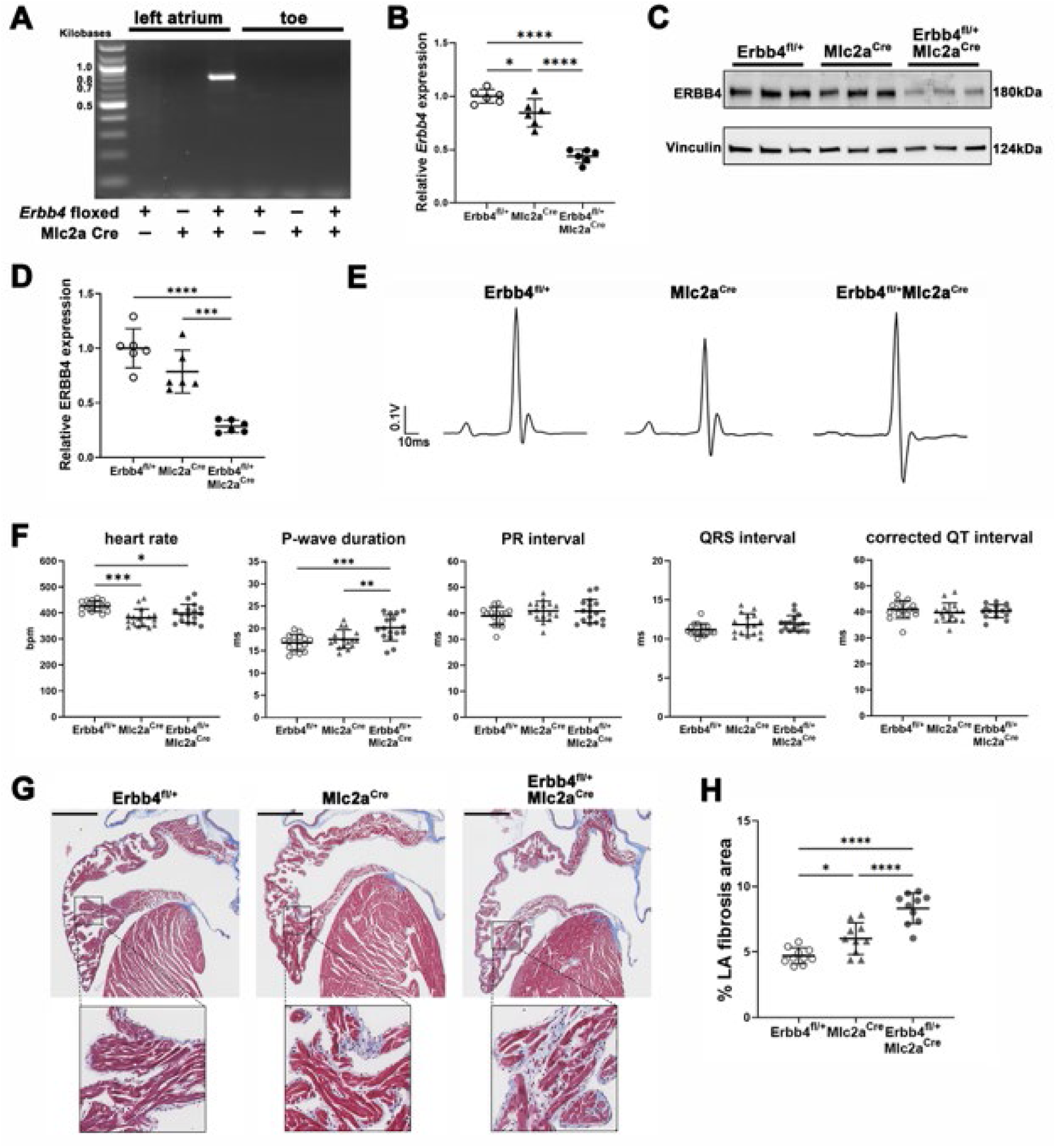
Cardiomyocyte-specific *Erbb4* reduction leads to atrial electrical and structural remodeling. **A**, Polymerase chain reaction (PCR) assay of DNA from left atrium (LA) and toe confirming targeted *Erbb4* exon 2 deletion in the atria of *Erbb4^fl/+^*Mlc2a^Cre^ mice. Band ∼756 bp represents recombined alleles. **B**, *Erbb4* expression in the LA by reverse transcription-quantitative PCR (RT-qPCR). Values are shown as mean±SD. \**P*<0.05, \*\*\*\**P*<0.0001 by one-way ANOVA followed by Tukey’s post-hoc test. n = 6 mice per group. **C**, Representative immunoblot assay for ERBB4. Molecular weight on immunoblots: ERBB4 ≈180 kDa and vinculin ≈124 kDa. **D**, Densitometric analysis of the immunoblot showing reduced ERBB4 expression in the LA of *Erbb4^fl/+^*Mlc2a^Cre^ mice. Data are shown as mean±SD. \*\*\**P*<0.001, \*\*\*\**P*<0.0001 by one-way ANOVA followed by Tukey’s post-hoc test. n = 6 mice per group. **E**, Representative surface electrocardiogram traces in sedated mice. **F**, Heart rate, P-wave duration, PR interval, QRS interval, and corrected QT interval across genotype groups. *Erbb4*^fl/+^Mlc2a^Cre^ mice exhibit prolonged P-wave duration compared to control mice. Data are shown as mean±SD. \**P*<0.05, \*\**P*<0.01, \*\*\**P*<0.001 by one-way ANOVA followed by Tukey’s post-hoc test. QRS interval was compared by Kruskal-Wallis test. n = 16 mice per group. **G**, Representative Masson’s trichrome-stained sections of the LA illustrating increased fibrosis in *Erbb4*^fl/+^Mlc2a^Cre^ mice. Scale bar 500 µm. Insets show magnified views indicated by black rectangles. **H**, Quantitative comparison of fibrosis area. The whole left atrial appendage was analyzed by ImageJ. Data are shown as mean±SD. \**P*<0.05, \*\*\*\**P*<0.0001 by one-way ANOVA followed by Tukey’s post-hoc test. n = 10 mice per group.

Surface electrocardiography was performed in the mouse model at 4 months of age (Figure 2E, F). Heart rates were lower in mice harboring the Mlc2a^Cre^ allele compared to those without the Cre allele (*Erbb4*^fl/+^). *Erbb4*^fl/+^Mlc2a^Cre^ mice had significantly longer P-wave duration compared to littermate controls (20.1±2.9 ms versus 16.7±1.8 ms, *Erbb4*^fl/+^, *P*=0.0005; versus 17.6±2.0 ms, Mlc2a^Cre^, *P*=0.0087). Other electrocardiographic parameters, including PR interval, QRS interval, and QT interval, were comparable between *Erbb4*^fl/+^Mlc2a^Cre^ and control mice. In addition to the standard parameters, we evaluated the cardiac electrical axis, which represents the overall direction of ventricular depolarization (Supplemental Figure S2A, B).^25,26^ *Erbb4*^fl/+^Mlc2a^Cre^ mice showed a shift in the QRS axis (68±13° versus 50±4°, *Erbb4*^fl/+^, *P*<0.0001; versus 59±11°, Mlc2a^Cre^, *P*=0.0350), suggesting a potential ventricular conduction abnormality in *Erbb4*^fl/+^Mlc2a^Cre^ mice. Transthoracic echocardiography was also performed to assess gross structural and functional changes between *Erbb4*^fl/+^Mlc2a^Cre^ and control mice (Supplemental Figure S2C). There were no significant differences in internal diameter of the LA, left ventricular contractile function measured by ejection fraction, right ventricular (RV) diameter, or RV systolic output assessed by pulmonary artery blood flow between *Erbb4*^fl/+^Mlc2a^Cre^ and each control group. Taken together, these findings demonstrate that selective *Erbb4* reduction in cardiomyocytes results in slowed atrial conduction in the absence of changes in ventricular function assessed by echocardiography.

Increased atrial fibrosis is a hallmark of atrial structural remodeling that contributes to atrial conduction disease and is associated with the persistence and recurrence of AF. To investigate whether structural remodeling underlies the observed conduction delay, we analyzed left atrial fibrosis using Masson’s trichrome staining in the mouse model. *Erbb4*^fl/+^Mlc2a^Cre^ LA showed an increased area of fibrosis compared to littermate controls, confirming the presence of structural remodeling in the LA of *Erbb4*^fl/+^Mlc2a^Cre^ mice (Figure 2G, H).

### Transcriptomic analysis in adult Erbb4 haploinsufficient mice

RNA-seq of left atrial tissue from *Erbb4*^fl/+^Mlc2a^Cre^ and control mice revealed distinct transcriptomic profiles. Principal component analysis of gene expression illustrates segregation of samples by genotype (Figure 3A). To detect DEGs specific to *Erbb4* deficiency and eliminate potential confounding by introduction of Cre recombinase, we individually compared *Erbb4*^fl/+^Mlc2a^Cre^ versus *Erbb4*^fl/+^ and *Erbb4*^fl/+^Mlc2a^Cre^ versus Mlc2a^Cre^ samples using DESeq2. Analysis of DEGs for each comparison using an adjusted *P* value cutoff (false discovery rate [FDR] < 0.05) is shown in Figures 3B and 3C. We identified genes that were differentially expressed in both individual comparisons, yielding a subset of 363 genes specific to *Erbb4* deficiency (Figure 3D). Within this subset, all but one gene, *Pam,* showed concordant directionality of expression change in both comparisons; *Pam* was therefore excluded from subsequent functional analysis (Pearson correlation coefficient R=0.933, *P*<0.0001; Figure 3E). Functional enrichment analysis was performed using Enrichr.^27^ Referencing GO biological processes, upregulated DEGs were significantly associated with regulation of ECM organization and apoptosis (Figure 3F, Supplemental Table S2A). In contrast, downregulated DEGs were involved in mitochondrial function and metabolism (Figure 3G, Supplemental Table S2B). Reactome pathway analysis revealed that upregulated DEGs were enriched for ECM organization and hemostasis (Figure 3H, Supplemental Table S2C), and downregulated DEGs were involved in pathways related to mitochondrial function, metabolism and cellular response to stress (Figure 3I, Supplemental Table S2D). Together, these results suggest that ERBB4 loss in cardiomyocytes disrupts transcriptional programs responsible for maintaining normal metabolic and structural properties in the atria.

**Figure 3.**
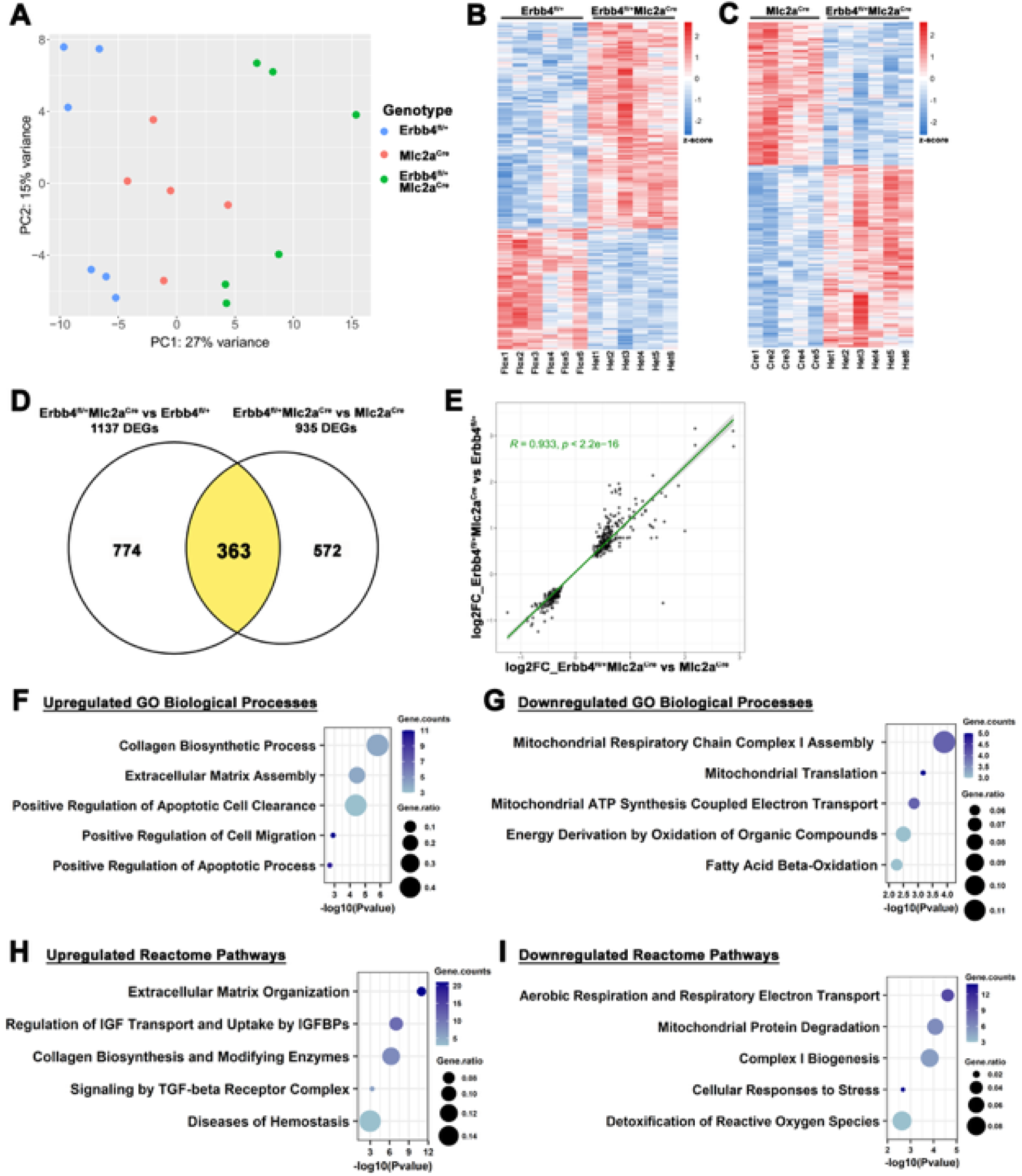
Left atrial transcriptome in ERBB4 haploinsufficient mice reveals gene expression changes related to cardiac pathophysiology. **A**, Principal component analysis plot of gene expression across three experimental genotypes. **B,** Heatmap of 1137 differentially expressed genes (DEGs) in *Erbb4^fl/+^*Mlc2a^Cre^ (n = 6) versus *Erbb4^fl/+^* (n = 6) left atria (LA). **C,** Heatmap of 935 DEGs in *Erbb4^fl/+^*Mlc2a^Cre^ (n = 6) versus Mlc2a^Cre^ (n = 5) LA. Adjusted *P* value cutoff for DEGs set at <0.05. **D**, Venn diagram showing overlap of DEGs from panels B and C; genes specific to *Erbb4* deficiency are represented by the overlapping 363 genes. **E**, Scatter plot of Pearson’s correlation of log_2_ fold change (FC) for genes specific to *Erbb4* deficiency across the two comparisons in panels B and C, resulting in 362 concordantly dysregulated genes. **F–I**, Dot plots of representative significantly upregulated and downregulated GO biological processes (F, G) and Reactome pathways (H, I) identified by Enrichr. Gene count is the number of input DEGs annotated within a GO term or Reactome pathway. Gene ratio is the proportion of the input DEGs to all annotated genes of each term. IGF, insulin-like growth factor; IGFBPs, IGF binding proteins; TGF, transforming growth factor.

NRG1/ERBB4 signaling is known to activate Ras/MAPK, PI3K/Akt, and Src/FAK pathways in cardiac myocytes.^28–31^ We therefore asked whether ERBB4 deficiency alters these downstream signals by assessing ERK and Akt phosphorylation in left atrial tissue from *Erbb4*^fl/+^Mlc2a^Cre^ and control mice. As canonical downstream effectors of Ras/MAPK signaling, phospho-ERK1/2 levels relative to total ERK were reduced in the LA of *Erbb4* haploinsufficient mice (Supplemental Figure S4). ERK1/2 coordinates multiple downstream effectors involved in mitochondrial dynamics, metabolic reprogramming, and stress adaptation,^32,33^ and reduced ERK1/2 activity likely contributes to the transcriptomic suppression of cellular stress-response pathways observed in *Erbb4*-deficient atria (Figure 3I and Supplemental Table S2D). This interpretation is consistent with prior evidence that cardiac ERK1/2 phosphorylates cytoplasmic and nuclear targets to modulate stress-responsive gene expression,^34^ and that genetic inhibition of ERK1/2 in cardiomyocytes predisposes the myocardium to decompensation and myocyte death under pathologic stress.^33^

Because AF GWAS have identified multiple candidate risk genes including *ERBB4*, we sought to determine if any AF-risk genes are differentially expressed in *Erbb4*^fl/+^Mlc2a^Cre^ left atria.^35^ Of the 362 DEGs in *Erbb4*^fl/+^Mlc2a^Cre^ left atria, two genes, *Gja5* and *Xpo7*, were shared with AF-GWAS risk loci. RT-qPCR confirmed that both *Gja5*, which encodes the high conductance gap junction protein Connexin 40, and *Xpo7*, which encodes the nuclear transport receptor Exportin-7, were downregulated in *Erbb4*^fl/+^Mlc2a^Cre^ mice compared to controls (Supplemental Figure S3A, B).

### ERBB4-associated gene networks are conserved between mouse and human

To investigate how gene expression profiles of *ERBB4* deficiency compare in the human LA, we utilized human left atrial RNA-seq data from the Cleveland Clinic Biobank.^20^ For cross-species analyses, mouse ERBB4-dependent genes were defined using DESeq2 with FDR < 0.1 as the initial subset. There were 760 DEGs in *Erbb4*^fl/+^Mlc2a^Cre^ left atria, of which 660 human orthologous genes could be assigned (Supplemental Figure S5A). Of the 660 orthologs, 646 were detectable in the left atrial RNA-seq dataset. Linear regression analysis between *ERBB4* and the orthologous transcripts identified that 534 (81%) of these orthologs demonstrated an association with *ERBB4* expression. Of these, 86% of the candidate genes behaved in a manner consistent with our murine model (460 orthologs, Supplemental Figure S5B)– that is, orthologs positively associated with *ERBB4* in the human LA (regression coefficient > 0 on the y-axis) were downregulated in *Erbb4*^fl/+^Mlc2a^Cre^ mice (log2FoldChange < 0 on the x-axis) and vice versa. Functional enrichment analysis of these 460 candidate genes using Enrichr demonstrated that genes positively associated with *ERBB4* were linked to GO biological processes including cardiac muscle cell contraction and conduction and mitochondrial function (Supplemental Figure S5C, Supplemental Table S3A). Conversely, genes negatively associated with *ERBB4* were linked to GO biological processes regulating ECM organization, apoptosis, inflammation, and coagulation (Supplemental Figure S5D, Supplemental Table S3B). Reactome pathway analysis showed that positively *ERBB4*-associated genes were primarily linked to mitochondrial function and metabolism (Supplemental Figure S5E, Supplemental Table S3C), whereas negatively *ERBB4*-associated genes were enriched for pathways involved in ECM organization, hemostasis, and inflammation (Supplemental Figure S5F, Supplemental Table S3D).

To validate these findings in a second human cohort, we utilized the open-access GTEx database containing RNA-seq data from right atrial appendages of 461 human subjects. Overall, *ERBB4*-correlated gene expression profiles and pathway enrichments in the GTEx right atrial dataset were highly consistent with those observed in the Cleveland Clinic Biobank dataset (Figure 4). Of the 660 mouse *Erbb4*-dependent orthologs, 649 were expressed in the GTEx dataset (Figure 4A). Correlation analysis revealed that 80% (517 genes) of these orthologs exhibited a significant correlation with *ERBB4* (FDR < 0.05). Of these, 83% of the *ERBB4*-correlated genes behaved in a manner consistent with our murine model (428 orthologs, Figure 4B). Further, we performed functional enrichment analysis using Enrichr to explore pathway-level associations. This analysis demonstrated that genes positively correlated with *ERBB4* were associated with mitochondrial function, including fatty acid β-oxidation and respiratory electron transport chain (ETC) activity, in GO biological processes (Figure 4C, Supplemental Table S4A). Conversely, genes negatively correlated with *ERBB4* were linked to ECM assembly, apoptosis, and inflammation (Figure 4D, Supplemental Table S4B). Reactome pathway analysis similarly showed that positively *ERBB4*-correlated genes were primarily linked to mitochondrial ETC and fatty acid β-oxidation (Supplemental Figure S6A, Supplemental Table S4C), whereas negatively *ERBB4*-correlated genes were enriched for pathways involved in ECM organization, hemostasis, and inflammation (Supplemental Figure S6B, Supplemental Table S4D). These findings support the hypothesis that *ERBB4* regulates structural and functional pathways in both mouse and human adult atrial tissue.

**Figure 4.**
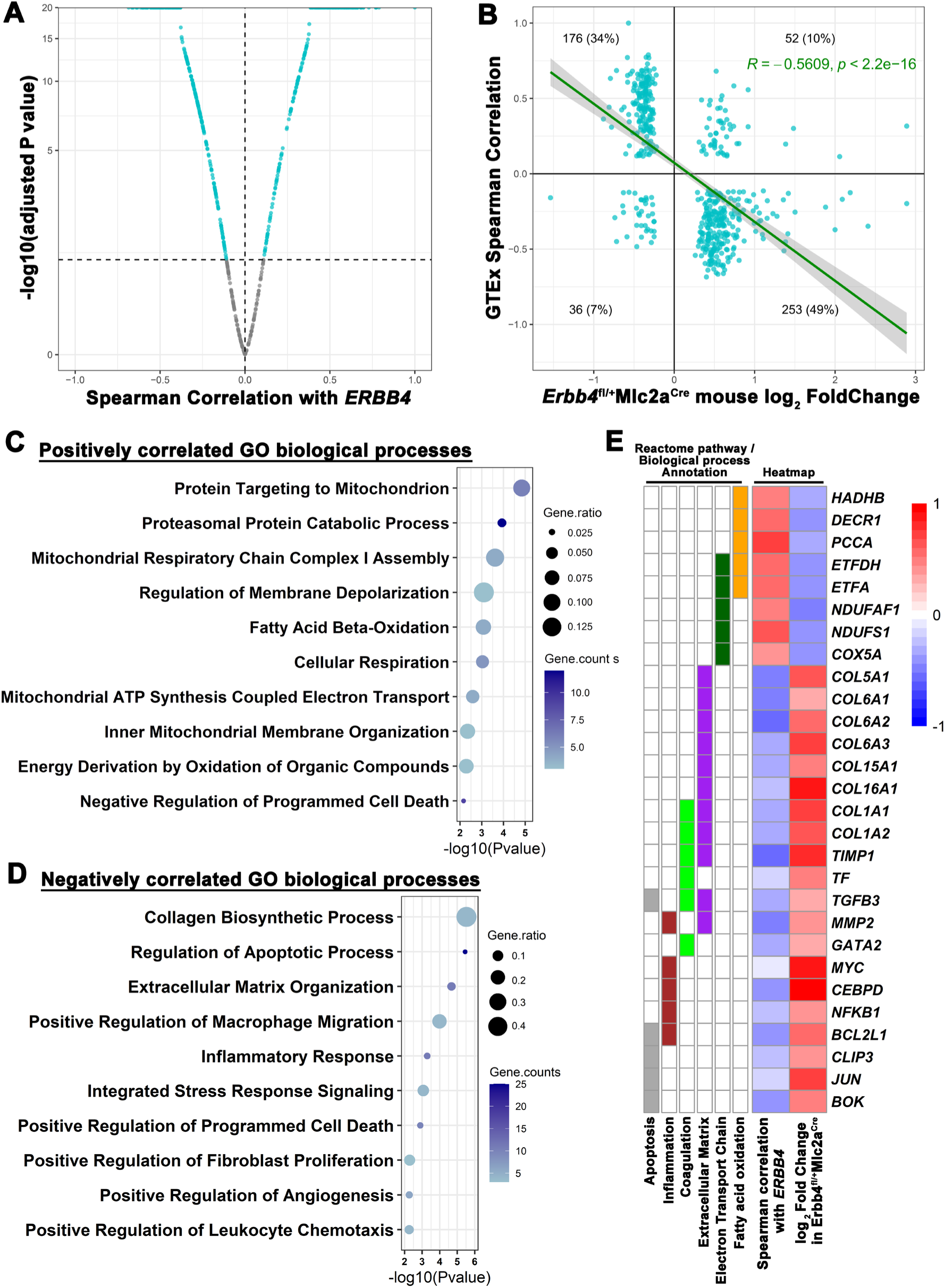
*ERBB4-*correlated gene expression patterns in human atria resemble those in the *Erbb4* haploinsufficiency mouse model. **A**, Volcano plot of Spearman correlation between *Erbb4* haploinsufficient mice orthologs and *ERBB4* expression levels in the GTEx right atrial appendage dataset. Each dot represents one of the 649 queried orthologs. Cyan dots indicate 517 orthologs significantly correlated with *ERBB4* (adjusted *P* value <0.05). **B**, Scatter plot showing the Pearson correlation (shown in green) between GTEx *ERBB4*-correlated human orthologs and differentially expressed genes in *Erbb4* haploinsufficient mice. A total of 429 orthologs show concordant directionality with *ERBB4*. **C and D**, Dot plots of representative functional enrichment analysis of the 429 concordant genes. Genes positively correlated with *ERBB4* in GTEx and upregulated in mouse are shown in (C); genes negatively correlated and downregulated are shown in (D). Gene ratio indicates the proportion of DEGs in all annotated genes in each GO term. **E**, Heatmap showing the correlation between *ERBB4* and individual genes in GTEx (left column) and log_2_ fold change in *Erbb4* haploinsufficient mice (right column). Color intensity represents the magnitude of correlation or log_2_ fold change. Red indicates positive correlation/upregulation, and blue indicates negative correlation/downregulation. Annotated Reactome pathways/GO biological processes of interest are shown on the left side of the heatmap with filled colors.

*ERBB4*-associated genes contributing to myocyte metabolism and atrial remodeling pathways are highlighted in Figure 4E. Key genes within fatty acid oxidation pathways that are positively correlated with *ERBB4* expression include *HADHB (*encoding β-subunit of the mitochondrial trifunctional protein), *DECR1* (encoding 2,4-dienoyl-CoA reductase), *ETFDH* (electron transfer flavoprotein dehydrogenase), and *ETFA* (encoding α*-*subunit of the electron transfer flavoprotein). We also identified key genes within ECM remodeling, prothrombotic, and proinflammatory pathways that are negatively correlated with *ERBB4* expression levels. The panel of ERBB4-regulated genes includes numerous collagen genes (*COL1A1*, *COL1A2*, *COL5A1*, *COL6A1*, *COL6A2*, *COL6A3*, *COL15A1*, *COL16A1*) and, notably, *TGFB3* (transforming growth factor β-3)*, MMP2* (matrix metalloproteinase-2)*, TIMP1* (tissue inhibitor of metalloproteinase-1), and *CEBPD* (CCAAT/enhancer binding protein delta).

Lastly, we assessed the relationship between AF-risk genes identified in our mouse model and *ERBB4* expression level in human left atria. *ERBB4* expression levels positively correlated with *GJA5* (Pearson correlation coefficient *R*=0.3492, *P*<0.0001) and *XPO7* (Pearson correlation coefficient *R*=0.5783, *P*<0.0001) (Supplemental Figure S3C), suggesting a conserved relationship between ERBB4 expression and AF-related gene expression in mouse and human left atria.

### Erbb4 deficiency in cardiomyocytes affects mitochondrial function

To validate transcriptomic changes and further characterize mitochondrial dysfunction, we performed immunoblotting on left atrial homogenates from *Erbb4*^fl/+^Mlc2a^Cre^ and control mice. Oxidative phosphorylation (Oxphos) is the most efficient way for the heart to produce energy (adenosine triphosphate, ATP), utilizing protein complexes I-V in the inner mitochondrial membrane (Figure 5A). We found significantly reduced levels of protein complexes III (CIII: UQCRC2, ubiquinol-cytochrome c reductase core protein II) and V (CV: ATP5A, alpha subunit of mitochondrial ATP synthase) in the *Erbb4*^fl/+^Mlc2a^Cre^ LA compared to controls (Figures 5B, C). We also found downregulation of HADHB, ETFDH and NDUFAF1 (NADH:ubiquinone oxidoreductase complex assembly factor 1) in *Erbb4*^fl/+^Mlc2a^Cre^ LA compared to controls, verifying transcriptomic results. Together, these data suggest a role for ERBB4 in maintaining effective cardiac mitochondrial metabolism.

**Figure 5.**
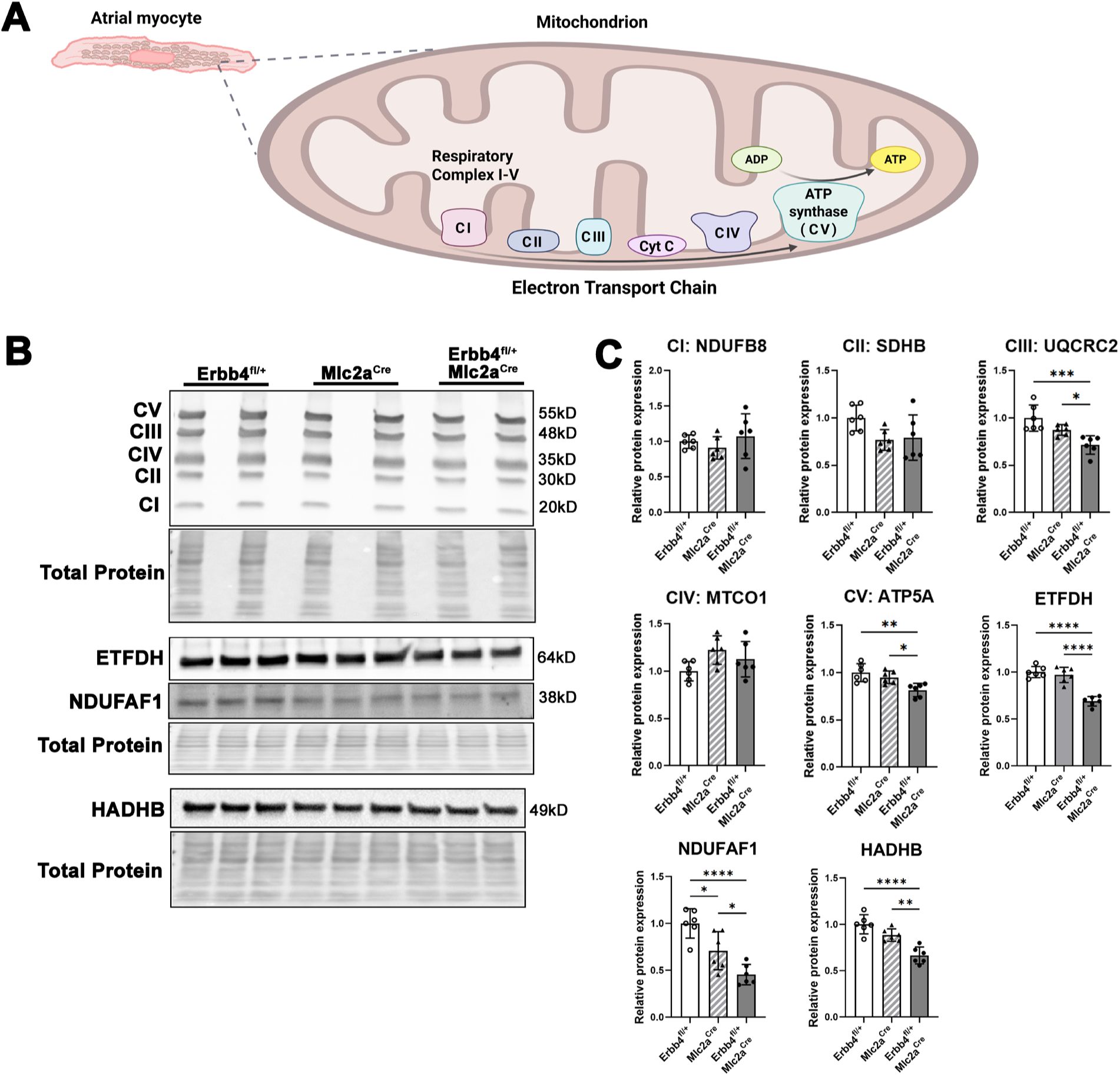
Reduction of ERBB4 in atrial myocytes alters mitochondrial protein expression. **A**, Schematic of mitochondrial electron transport chain complexes in atrial cardiomyocytes. Image created using biorender.com. CI, complex I; CII, complex II; CIII, complex III; CIV, complex IV; CV, complex V (ATP synthase); Cyt C, cytochrome C; ADP, adenosine 5’-diphosphate; ATP, adenosine 5’-triphosphate. **B**, Representative immunoblot of whole left atrial lysates of Erbb4^fl/+^Mlc2a^Cre^ and control mice. Molecular weights were estimated based on migration relative to the protein ladder. Total protein images were uniformly resized for display. **C**, Densitometric quantification showing reduced expression of mitochondrial respiratory and metabolic proteins. Target proteins are normalized to total protein. Data are shown as mean±SD. \**P*<0.05, \*\**P*<0.01, \*\*\**P*<0.001, \*\*\*\**P*<0.0001 by one-way ANOVA followed by Tukey’s post-hoc test. n = 6 mice per group.

Mitochondria produce reactive oxygen species (ROS) as a natural by-product of aerobic metabolism during ETC activity for ATP synthesis.^36^ Mitochondria have been identified as a predominant source of cellular superoxide, which is the main ROS species in the heart. Excessive ROS has been linked to AF in human and animal studies.^37,38^ MitoSOX is a fluorogenic dye that is taken up by mitochondria and oxidized by superoxide to visualize mitochondrial superoxide production. MitoSOX staining of left atrial myocytes revealed increased mitochondrial superoxide in *Erbb4*^fl/+^Mlc2a^Cre^ mice compared to controls (Figures 6A, B), indicating dysregulated mitochondrial ROS production in atrial myocytes. Notably, transmission electron microscopy revealed an altered number of mitochondria in left atrial cardiomyocytes in both *Erbb4*^fl/+^Mlc2a^Cre^ and Mlc2a^Cre^ mice (Supplemental Figure S8), suggesting that Cre knock-in insertion may lead to decreased mitochondrial abundance. However, mitochondrial dysfunction, as evidenced by increased ROS production, was specific to the *Erbb4* haploinsufficient mice.

**Figure 6.**
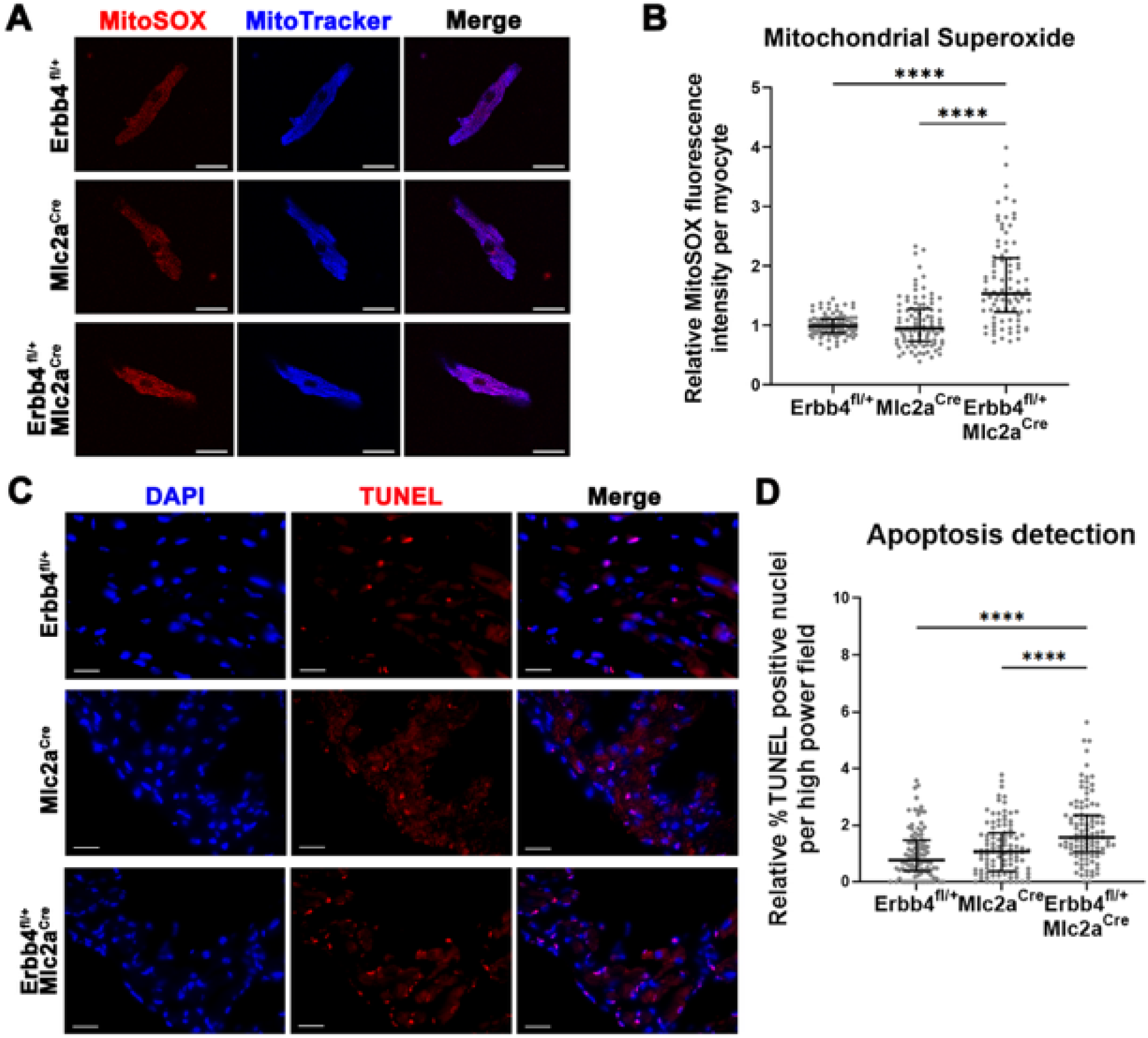
Cardiac ERBB4 haploinsufficiency leads to pathological remodeling in the mouse left atrium. **A**, Representative confocal images of MitoSOX (red) and MitoTracker (blue) staining for mitochondrial superoxide assay in LA myocytes. Scale bar 25 µm. **B**, Quantitative comparison of relative MitoSOX fluorescence intensity analyzed by ImageJ. Values are shown as median with interquartile range. \*\*\*\**P*<0.0001 by Kruskal-Wallis test followed by Dunn’s post-hoc test. 91–100 myocytes analyzed from 4 mice per group. **C**, Representative images of terminal deoxynucleotidyl transferase dUTP nick end labeling (TUNEL, red) and 4′,6-diamidino-2-phenylindole (DAPI, blue) staining of left atrial (LA) sections. Scale bar 20 µm. **D**, Quantitative comparison of the percentage of TUNEL positive nuclei per high power field (HPF). Whole LA area was visualized and analyzed. Each dot represents one HPF image. Values are shown as median with interquartile range. \*\*\*\**P*<0.0001 by Kruskal-Wallis test followed by Dunn’s post-hoc test. Total 97–103 HPF images were collected from 4 mice per group.

### Increased apoptosis in Erbb4-deficient atria

Cardiomyocyte apoptosis has been implicated as a key cellular process in the pathogenesis of cardiomyopathy^39,40^ and identified as a key dysregulated pathway in diseased atria in our mouse models (Figure 1C,D and Figure 3F). Moreover, apoptosis is a known downstream consequence of increased mitochondrial ROS.^41–43^ To quantify apoptosis in the LA, we performed histological assessment by terminal deoxynucleotidyl transferase dUTP nick end labeling (TUNEL) staining. We observed an increase in the percentage of TUNEL-positive nuclei as a proportion of total LA area in *Erbb4*^fl/+^Mlc2a^Cre^ mice compared to controls (Figures 6C, D, Supplemental Figure S7). Activation of caspases, a family of cysteine-aspartic acid proteases, plays a central role in the cascade of apoptotic cell death, as distinct from necrotic cell death. Previous studies have demonstrated that myocardial apoptosis is accompanied by increased cleavage of caspase-3 in rodent cardiomyopathy models.^44,45^ We measured cleaved caspase-3 by immunoblotting and found increased cleaved caspase-3 expression in *Erbb4^fl/+^*Mlc2a^Cre^ LA compared to that in control mice (Supplemental Figure S4A, B).

## Discussion

Genetic studies have identified variants near *ERBB4* associated with the incidence and recurrence of AF,^1,46^ highlighting ERBB4 as a candidate mediator of AF risk. This study shows that ERBB4 plays a critical role in maintaining normal atrial physiology in adult hearts. Using a mouse model of cardiomyocyte-specific *Erbb4* deficiency, we observed atrial conduction disease, increased left atrial fibrosis, apoptosis, and altered cellular metabolism, creating an atrial substrate primed for arrhythmia. In human atrial RNA-seq datasets, orthologous genes identified as *Erbb4*-dependent in our mouse model showed concordant expression with *ERBB4* and were enriched for functional pathways similar to those altered in *Erbb4* haploinsufficient mouse atria.

An important finding in our study is that ERBB4 deficiency disrupts pathways related to fatty acid oxidation and mitochondrial ETC, mirroring observations in patients with permanent AF. In right atrial tissue from patients with permanent AF, genes involved in fatty acid oxidation are downregulated along with a corresponding reduction in *ERBB4* expression.^47^ In normal fasting human hearts, fatty acid oxidation provides up to 85% of ATP production.^48^ In the *Erbb4*^fl/+^Mlc2a^Cre^ mouse LA, downregulation of fatty acid oxidation pathways included genes such as *HADHB*, *ETFDH*, *ETFA*, and *DECR1*. *HADHB* is essential for long-chain fatty acid β-oxidation,^49^ and homozygous loss-of-function mutations in *HADHB* are associated with neonatal lethality from severe cardiomyopathy.^49^ Mutation in either *ETFDH* or *ETFA* causes electron transfer flavoprotein deficiency (clinically known as multiple acyl-CoA dehydrogenase deficiency/glutaric aciduria type II), a lipid storage myopathy due to impaired fatty acid oxidation.^50^ This can present as a severe neonatal-onset form with hypertrophic cardiomyopathy and sudden cardiac death^51^ or a milder late-onset form associated with supraventricular arrhythmias.^52^ *DECR1* is an auxiliary enzyme in mitochondrial β-oxidation of unsaturated fatty acids,^53^ and *Decr1* knockout mice display tissue accumulation of unsaturated fatty acids.^53^ Conversely, *Decr1* deficiency can ameliorate cardiac fibrosis and myopathy in diabetic models by limiting excessive fatty acid oxidation,^54,55^ highlighting the need for tight regulation of fatty acid oxidation.

Our RNA-seq analysis also reveals downregulation of respiratory ETC pathways in *Erbb4*^fl/+^Mlc2a^Cre^ atria, aligning with transcriptomic and functional studies in patients with persistent AF. A study comparing patients with AF to comorbidity-matched non-AF patients demonstrated decreased ETC activity and increased mitochondrial ROS generation in atrial tissue samples.^56^ Reduced ETC activity in human atria has been attributed in part to decreased activity of mitochondrial respiratory complex I, which oxidizes NADH for ATP production.^56^ In our datasets, mitochondrial complex proteins involved in normal ETC assembly and function were altered, including UQCRC2, ATP5A, and NDUFAF1. Loss-of-function mutation of *NDUFAF1* is associated with human cardiomyopathic states, likely driven by energy deficiency and disrupted mitochondrial homeostasis in cardiomyocytes.^57^ Although loss-of-function variants in *UQCRC2* and ATP5A (ATP5F1A) have not been clearly linked to cardiomyopathy, pathogenic mutations in other ETC subunits frequently cause mitochondrial cardiomyopathy, supporting a role for ERBB4-dependent ETC dysregulation in atrial metabolic remodeling in persistent AF. Taken together, our data support a role for ERBB4 in regulating genes critical for normal fatty acid oxidation and mitochondrial ETC activity, thereby providing a mechanistic link between altered ERBB4 levels due to genetic predisposition or pressure overload states and atrial metabolic remodeling.

Increased ROS generation can trigger cellular autophagy and apoptosis.^41–43^ Autophagy is the lysosome-dependent intracellular degradation system by which harmful cytoplasmic components including damaged mitochondria are eliminated to maintain cellular homeostasis. Apoptosis, in contrast, is the process of programmed cell death characterized by caspase activation and DNA fragmentation in response to environmental or internal cues. In both animal and human studies, higher levels of apoptosis have been observed in failing hearts.^58^ It has been also suggested that autophagy and apoptosis demonstrate crosstalk, and inactivation of autophagy may promote apoptosis.^59^ In our RNA-seq data, pathways associated with autophagy were downregulated and those associated with apoptosis upregulated in *Erbb4* haploinsufficient mice (Supplemental Table S2B). The combination of increased ROS and dysregulation of cellular autophagy provides a potential mechanistic explanation for the higher rates of cardiomyocyte apoptosis observed in the mutant cohort.

In addition to regulating atrial metabolism, our work also implicates ERBB4 in regulating genes involved in atrial conduction, inflammation, fibrosis, and coagulation-key pathways dysregulated in AF. *GJA5/*Connexin 40 enables rapid impulse propagation between atrial myocytes, and mutations in *GJA5* are associated with AF.^60^ By contrast, little is known about the role of *XPO7* in cardiac physiology and pathology, representing an opportunity for discovery. Beyond AF-GWAS-identified risk genes, genes involved in pathological atrial remodeling and particularly those driving pro-inflammatory (*MMP2*, *CEBPD*), pro-fibrotic (collagen genes, *TGFB3*, *MMP2, TIMP1*), and prothrombotic (*COL1A1*, *COL1A2*, *TGFB3*, *TIMP1*) processes were found to be dysregulated with *Erbb4* deficiency. *TGFB3* plays a key role in fibrotic signaling, with gain-of-function mutations linked to arrhythmogenic cardiomyopathy.^61^ MMP2 contributes to atrial remodeling and heart failure,^62,63^ with elevated plasma levels linked to AF.^64^ MMP2 acts as a mediator of cardiac proinflammatory signaling^65^ and ECM remodeling.^62,63^ Elevated serum TIMP1 is associated with atrial remodeling, AF recurrence after catheter ablation, and a prothrombotic state.^66–68^ *CEBPD* encodes CCAAT/enhancer-binding protein delta, a transcription factor that induces inflammatory cytokines^69^ and promotes cardiac fibrosis via upregulation of connective tissue growth in cardiomyocytes.^70^ Together, our data implicate diminished ERBB4 signaling as a key mediator of metabolic, pro-inflammatory, profibrotic, and prothrombotic remodeling in the persistent AF atrium.

Several observations from this work warrant further investigation. Our study utilized bulk RNA-seq approaches, which, while providing insights into the overall atrial tissue environment, do not resolve cell type-specific transcriptional programs or cell-to-cell communication. In addition, the cardiomyocyte-selective Cre driver utilized in this study is constitutive. Although it provides an excellent genetic model of *Erbb4* haploinsufficiency, it does not adequately model acquired *Erbb4* deficiency. Our laboratory has generated inducible cardiomyocyte-selective *Erbb4* deficiency models and comprehensive physiological and molecular studies, including single-nucleus RNA-seq analysis, are currently underway. These studies will explore how acute deletion of *Erbb4* in cardiomyocytes secondarily remodels neighboring cell populations such as fibroblasts, macrophages, and other atrial cell types.

In conclusion, our study identifies ERBB4 as an essential signal transducer for maintaining atrial metabolic, electrical, and structural integrity. ERBB4 maintains atrial health by preserving normal cardiomyocyte metabolism while preventing apoptosis, a procoagulant state, and fibrosis. Targeting ERBB4-dependent pathways is currently being explored^71^ and may represent a potential therapeutic strategy for preventing the development and progression of pathological atrial remodeling that leads to AF.

## Data Availability

All data referred to in the manuscript are available within the article, its Supplemental Material, or from publicly accessible repositories.

## Acknowledgements

We are grateful to Experimental Pathology Research Laboratory at NYU Langone’s Laura and Isaac Perlmutter Cancer Center for their technical assistance, to Dr. Alice Liang and her team in NYU Langone Health Microscopy Lab for their assistance in the acquisition and analysis of electron microscopy images, to Dr. Michael Cammer for his comments and suggestions regarding immunohistochemical staining of cardiac sections and analysis of electron microscopy images, and the Genome Technology Center (RRID:SCR_017929) at NYU Langone for their assistance in the generation of mouse RNA-seq data.

## Supplemental Material

Supplemental Methods

Supplemental Figures S1–8

Supplemental Files S1–4

## Non-standard Abbreviations and Acronyms

AF: atrial fibrillation
AngII: angiotensin II
ATP: adenosine triphosphate
Cre: cre recombinase
DEGs: differentially expressed genes
ECG: electrocardiography
ECM: extracellular matrix
ERK: extracellular signal-regulated kinase
ETC: electron transport chain
FAK: focal Adhesion Kinase
GO: gene ontology
GTEx: genotype-tissue expression project
GWAS: genome-wide association studies
LA: left atrium
LV: left ventricular
MAPK: mitogen-activated protein kinase
MLC2a: myosin regulatory light chain 2, atrial isoform
MLC2v: myosin regulatory light chain 2, ventricular isoform
PI3K: phosphatidylinositol 3 kinase
RNA-seq: RNA sequencing
ROS: reactive oxygen species
RT-qPCR: reverse transcription-quantitative polymerase chain reaction
RV: right ventricular
SRC: c-Src proto-oncogene
TAC: transverse aortic constriction

