## Supplemental methods and figures for "ERBB4 deficiency promotes atrial myopathy underlying the atrial fibrillation substrate"


**SUPPLEMENTAL MATERIAL contains:**

**Detailed Methods**

**Supplemental Figures S1 – S8**

**Supplemental Tables S1 – S4**

### Detailed Methods

#### Experimental Animals

All animal experimental procedures were approved by the Institutional Animal Care and Use Committee of New York University Grossman School of Medicine (protocol#PROTO201900150), and the animals received humane care in accordance with the US National Institutes of Health Guide for the Care and Use of Laboratory Animals.

#### Cardiomyocyte-Specific *Erbb4* Heterozygous Mice

*Erbb4*$^{fl/+}$ mice carrying a loxP-flanked *Erbb4* allele (B6;129-Erbb4tm1Fej/Mmucd, Mutant Mouse Resource & Research Centers, #010439-UCD) were crossed with Mlc2a$^{Cre}$ mice carrying Cre recombinase under the control of myosin regulatory light chain 2 (MLC2a) promotor (*Myl7*$^{tm1(cre)Krc}$)[1] in C57BL/6J background to generate mice with cardiomyocytes-specific heterozygous deletion of *Erbb4*. Cre-loxP-mediated *Erbb4* exon 2 excision was confirmed by polymerase chain reaction (PCR)-based methods using the following primers: forward, 5'-gcaccagcagaccactttct-3'; reverse, 5'-acgagggaatttcgacggat-3'. Both male and female *Erbb4*$^{fl/+}$Mlc2a$^{Cre}$ and littermate (*Erbb4*$^{fl/+}$ and Mlc2a$^{Cre}$) mice were used at 4 months of age in this study.

#### Surface Electrocardiography (ECG)

For surface ECG measurements, *Erbb4*$^{fl/+}$Mlc2a$^{Cre}$ and control (*Erbb4*$^{fl/+}$ and Mlc2a$^{Cre}$) mice were anesthetized with 1.5 % isoflurane using a nose cone. Mice (N=16 per group) were placed in supine position and warmed to maintain a stable body temperature at 37±0.5 °C. Mouse body temperature was monitored via a rectal probe during recordings. Surface ECG signals from standard bipolar limb leads I, II, and III were recorded for 3 minutes. Parameters from averaged ECG traces for at least 30 seconds on lead II were analyzed in blinded fashion using ECG Analysis module in LabChart Pro software (v8, ADInstruments Inc., CO, USA) according to the instructions. Measured parameters included heart rate, P-wave duration, PR interval, QRS interval, and QT interval in 16 animals per group. The P-wave duration included both the first positive and negative deflections from baseline. PR interval was measured from the onset of the P wave to the onset of the QRS complex. QRS interval was determined from the initial upward deflection from baseline after the P wave until the maximum deflection in the negative wavefront between S wave and the following positive wave right after the QRS complex representing early repolarization. QT intervals were corrected for RR intervals using Mitchell's formula.[2]

For calculation of frontal mean QRS axis, the average beat for lead I and II was computed and analyzed using LabChart software. Mean amplitude of R peaks and S peaks were detected automatically. The net QRS amplitudes in lead I and II were calculated as the difference between R and S wave amplitudes. Then QRS axis was calculated using atan2 formula, which accounts for the correct quadrant: $\theta = \text{atan2}(\text{lead II amplitude}, \text{lead I amplitude}) * (180/\pi)$ in individual mouse (n=16 per group). For visualization, the angle data were plotted with the axes oriented to align with the standard ECG convention, where 0° corresponds to the lead I direction, 90° points downward, and -90° points upward using RStudio.

### Transthoracic Echocardiography

A 30-Mhz linear array transducer and a Vevo F2 Imaging System (FUJIFILM VisualSonics, Inc., Toronto, Canada) were used for transthoracic echocardiographic recordings. *Erbb4*$^{fl/+}$Mlc2a$^{Cre}$ and control (*Erbb4*$^{fl/+}$ and Mlc2a$^{Cre}$) mice were anesthetized with 2 % isoflurane via a nose cone. Mice were positioned supine and warmed on a heated platform to maintain a stable body temperature at 37±0.5 °C. Mouse body temperature was monitored via a rectal probe during recordings. Parasternal long axis view in 2-dimensional B-mode and short axis view at the papillary muscle level in M-mode were obtained. The recordings were analyzed with VevoLAB analysis software (v3.2.0, Fujifilm VisualSonics, Inc.) according to the manufacturer's analysis guideline in a blind fashion. For left atrial diameter, distance from ventral (relative to anterior in human) to dorsal (relative to posterior in human) wall of the left atrium (LA) was measured at the aortic valve level at end-systole. For the left ventricular dimension and function, at least 3 consecutive M-mode cycles were analyzed then averaged. For the right ventricular internal dimension, distance from ventral free wall to interventricular septum of the right ventricle was measured at the level of the mitral valve annulus at end-diastole on a B-mode modified long axis view. To measure pulmonary artery velocity-time integral (VTI), pulse-wave doppler was obtained with the sample volume placed just proximal to the pulmonary artery. All measurements were averaged from at least 3 consecutive cardiac cycles.

**Histological assessment**

For histology, whole hearts were excised and rinsed in ice-cold phosphate buffered saline (PBS), then fixed in 4% paraformaldehyde (PFA) overnight at 4 °C. The samples were stored in 70% ethanol until processed for paraffin sectioning. Before embedding, some hearts were visualized for bright-field whole mount images using the Zeiss M2Bio microscope equipped with a Zeiss AxioCam Color camera interfaced with Zeiss Axiovision 2012 software. The fixed samples were sectioned at 5 μm-thickness. Staining with hematoxylin and eosin and Masson's trichrome were performed according to standard protocols at New York University Experimental Pathology Research Laboratory. All bright field images were acquired using BZ-X800 microscope (Keyence, Osaka, Japan). For quantification of atrial fibrosis, whole LA was analyzed using ImageJ Fiji software (1.54f, National Institutes of Health, USA) in a blind manner.[3] First, a Colour Deconvolution plugin was used with a vector of Masson Trichrome. Then, a color threshold setting was applied on the Coulor-3 image (green). Lastly, the LA area was selected and measured for area fraction occupied by fibrosis.

**Immunoblot assay**

Left atrial appendages were collected from *Erbb4*$^{fl/+}$Mlc2a$^{Cre}$ and control mice. Samples were immediately cryopreserved in liquid nitrogen and stored at –80 °C before protein extraction. Samples were sonicated in RIPA lysis buffer (#89900, Thermo Fisher Scientific, Waltham, MA) containing 1 % of protease and phosphatase inhibitors (Thermo Fisher

Scientific). Protein concentration was determined using a Bradford protein assay with a spectrophotometer. Processed protein samples were loaded and run on 4−20% stain-free gel (Bio-rad) and transferred to a nitrocellulose membrane following a standard Western blot protocol (Bio-rad). The following antibodies were used: anti-ERBB4 (#4795; Cell Signaling Technology, Danvers, MA), anti-p44/42 MAPK (#9107, Cell Signaling Technology), anti-phospho-p44/42 MAPK (#9101, Cell Signaling Technology), anti-Akt (#2920, Cell Signaling Technology), anti-phospho-Akt (#4060, Cell Signaling Technology), anti-Vinculin (#26520-1-AP, Proteintech, Rosemont, IL), anti-cleaved caspase 3 (#9664, Cell Signaling Technology), anti-OxPhos cocktail (#45-8099, Thermo Fisher Scientific), anti-ETFDH (#11109-1-AP, Proteintech), anti-NDUFAF1 (#15181-1-AP, Proteintech), anti-HADHB (#48241, Cell Signaling Technology), Goat anti-Mouse IgG (#926-68070, LI-COR biotechnology, Lincoln, NE), and Goat anti-Rabbit IgG (#926-32211, LI-COR biotechnology). Antigen complexes were visualized with ChemiDoc MP Imaging system (BioRad) and band intensity was quantified using Image Studio Lite software (v5.5, LI-COR Biosciences). Vinculin was used as loading control for non-mitochondrial proteins, whereas mitochondrial proteins were normalized to total protein.

**Mouse RNA sequencing**

RNA sequencing (RNA-seq) was performed with total RNA extracted from atrial tissue at 4-months-old *Erbb4* deficient mouse models. Left atrial appendages were collected from 6 animals per group (*Erbb4*$^{fl/+}$, Mlc2a$^{Cre}$ and *Erbb4*$^{fl/+}$Mlc2a$^{Cre}$) as biological replicates. Total RNA was extracted using the RNeasy Plus Mini Kit (74134, Qiagen Inc., MD, USA). For each sample, isolated RNA was amplified, and sequencing libraries were prepared using the Low Input Clontech SMART-Seq RNA Kit (Nugen, CA, USA) with 50 bp paired-ended at 10 to 20 million reads per replicate on an Illumina NextSeq 6000 instrument. Sequencing reads were mapped to the reference genome (mm10) using the STAR aligner (v2.6.1d).[4] The read count tables were generated using FeatureCounts (v1.6.3)[5] and normalized based on their library size factors using DESeq2 (v1.42.1).[6] Using DESeq2 package, differential expression analysis was performed with Wald test and the *P* values were corrected by Benjamini-Hochberg method. To compare the level of similarity between the groups and their replicates, principal component analysis (PCA) was used. One $Mlc2a^{Cre}$ replicate was excluded from further analysis based on the PCA plot. Significant differentially expressed genes (DEGs) between $Erbb4^{fl/+}Mlc2a^{Cre}$ and control groups, were selected based on adjusted $P$ value cut-off of 0.05 (false discovery rate < 0.05). Shared DEGs between the two datasets: $Erbb4^{fl/+}Mlc2a^{Cre}$ versus $Erbb4^{fl/+}$ and $Erbb4^{fl/+}Mlc2a^{Cre}$ versus $Mlc2a^{Cre}$ were used for Pearson's correlation analysis to obtain the fundamentally altered DEGs by ERBB4 reduction. Functional enrichment analysis was performed with the common DEGs using an open online source Enrichr[7] to find pathways enriched from the Gene Ontology Biological Process and Reactome databases. Representative terms/pathways by $P$ value (< 0.05) were selected from each library. All statistical analyses and generating plots were performed in R environment (v4.0.3).[8] The RNA-seq dataset from adult *Erbb4* deficiency mouse model has been deposited into NCBI GEO database (GSE288616).

Other RNA-seq datasets from cardiac pressure overload mouse models induced by transverse aortic constriction (TAC) and angiotensin II infusion[9,10] were obtained (GSE322182) and reanalyzed using the same pipeline described above. To identify concordant transcriptional responses shared between the two models, overlapping DEGs were selected. For pathway analysis, the shared DEGs were ranked by the average log2 fold change across the TAC and angiotensin II datasets, and preranked gene set enrichment analysis (GSEA) was performed in R using the clusterProfiler package. GO Biological Process enrichment was assessed using Enrichr, and Hallmark pathway enrichment was performed using GSEA with MSigDB Hallmark gene sets obtained through msigdbr. Genes were mapped from mouse gene symbols to Entrez IDs using org.Mm.eg.db. Pathways with adjusted P < 0.05 were considered significantly enriched.

**Reverse transcription (RT)- quantitative PCR**

Left atrial appendage was collected from adult experimental mice and stored at −80 °C before RNA extraction. Total RNA was extracted using the RNeasy Plus Mini Kit (Qiagen

Inc.). For quantification of gene expression levels of interest, cDNA synthesis was performed using Maxima First Strand cDNA Synthesis Kit for RT-qPCR (K1641, Thermo Fisher Scientific, MA, USA), then qPCR was performed using Power SYBR Green PCR Master Mix (Thermo Fisher Scientific) on a StepOne Real-Time PCR System (Applied Biosystems). All primer pairs for qPCR were purchased from Origene (MD, USA) except *Erbb4* exon 2 specific primers: forward, 5'- cgagccttgcgcaaatacta-3'; reverse, 5'-acttctcggatagaccgcag-3', and used according to the manufacturer's protocol. Target gene expression was normalized to glyceraldehyde-3-phosphate dehydrogenase (gapdh), and fold changes were calculated using the comparative $2^{-\Delta\Delta CT}$ method.

**GTEx ERBB4 Correlation analysis**

Human right atrial RNA-seq data was downloaded from Genotype Tissue Expression (GTEx) portal on January 24th 2025.[11] Read counts were normalized using DESeq2 size factors, and log transformed via the "log1p" function in R. Spearman correlation coefficients between each gene and *ERBB4* expression were calculated using the "cor.test" function in R,  and resulting P values were adjusted for multiple testing using the Benjamini–Hochberg false discovery rate (FDR) procedure implemented in the "p.adjust" function. Mouse–human orthologs were defined in R using the MGI Mouse–Human homology report (DB.Class.Key–based grouping of genes), supplemented with Ensembl gene annotations to add missing orthologs, and then curated to yield a set of 661 genes, consisting primarily of one to one ortholog for cross-species analyses. The R package "fgsea" (v1.28)[12] was used to perform GSEA analysis after sorting GTEx genes by Spearman rho. *ERBB4*-correlated genes were selected based on adjusted *P* value cut-off of 0.05 (false discovery rate < 0.05). The GSEA results were summarized and visualized as dot plot in R. Positively and negatively *ERBB4*-correlated genes were analyzed using Enrichr for functional enrichment.[7]

**Human left atrial RNA-seq analysis**

Previously reported RNA-seq dataset from Cleveland Clinic Biobank were reanalyzed for this study. Detailed methods of generating sequencing data from left atrial appendage samples are described in Hsu, et al.[13] Human subjects' characteristics are also summarized in Yamaguchi, et al.[9] The RNA-seq data are deposited in the NCBI GEO database (GSE69890). Out of total 265 specimens, 14 donor samples were excluded due to missing data values. 251 subjects who underwent cardiac surgery were included for the analysis in this study. For comparison of the cardiac rhythm at the time of surgery, 7 subjects were excluded due to the lack of rhythm diagnosis.

For the cross-species analysis, the same subset of 661 orthologs described above was used.  Overlapping human orthologs in the left atrial RNA-seq dataset were analyzed for association between *ERBB4* and each transcript using simple linear regression with JMP software (JMP Statistical Discovery LLC, NC, USA). Estimate values were used as unstandardized regression coefficient to represent the direction and magnitude of associations. Prob>F values were used as $P$ values without adjustment for multiple comparisons.

**Terminal deoxynucleotidyl transferase dUTP Nick-End Labeling (TUNEL) staining on heart section**

For apoptotic cell detection in mouse LA samples, TUNEL staining was performed on paraffin-embedded heart sections at 5 µm-thickness using TUNEL assay kit (#ab66110, Abcam, MA, USA) according to the manufacture's protocol for IHC Detection. Briefly, paraffin sections were deparaffinized and rehydrated sequentially, then incubated with proteinase K solution for 5 minutes followed by refixation with 4% PFA. After washing steps, sections were stained with antibody solution containing terminal deoxynucleotidyl transferase enzyme and brominated deoxyuridine triphosphate nucleotide (Br-dUTP) for 60 min at 37 °C, then incubated with anti-BrdU-Red antibody solution for 30 min at room temperature. Stained sections were covered with a glass coverslip with an antifade mounting medium with DAPI (VECTASHIELD H-1200, Vector Laboratories, Inc., CA, USA), and visualized using BZ-X800 microscope (Keyence, Osaka, Japan). The BrdU fluorescent nucleus was counted by transferring DAPI-originated regions of interest (blue channel) to threshold BrdU-positive regions (red channel) using ImageJ Fiji (1.54f) and the built-in Analyze Particle function. The percentage of BrdU positive nuclei was calculated per image and compared between groups. Some slides were stained with anti-alpha actinin antibody (ab18061, Abcam) prior to TUNEL assay for left atrial tissue visualization. Co-stained sections were excluded from quantitative analysis.

**Measurement of reactive oxygen species (ROS) in left atrial myocytes**

*Erbb4*$^{fl/+}$Mlc2a$^{Cre}$ and control mice were injected with 125IU of heparin intraperitoneally and euthanized by inhalation of 100% $CO_2$. The hearts were surgically excised and cannulated via the aorta with 20G blunted-needle, then perfused with a digestion buffer containing (in mmol/l); 113 NaCl, 4.7 KCl, 1.2 $MgSO_4$, 0.6 $Na_2HPO_4$, 0.6 $KH_2PO_4$, 12 $NaHCO_3$, 10 $KHCO_3$, 10 HEPES and 30 Taurine, and enzymes; collagenase type II (29.9kU, Worthington, Lakewood, NJ, USA) and trypsin at a constant flow rate of 3 ml/min at 37°C. After digestion, LA was teased into small pieces with fine-tipped forceps, and gently triturated with a plastic Pasteur pipette. Then the cell suspension was filtered with a 100μm filter. The isolated cardiomyocytes were treated with $Ca^{2+}$ containing buffer, then plated on a glass-bottom dish coated with poly-D-lysine and laminin. Superoxide levels were assessed by incubating myocytes with mitochondria specific ROS indicator, MitoSOX Red (5μM, #M36007, ThermoFisher Scientific) in Hanks' balanced salt solution (HBSS) buffer for 15 minutes at 37°C. After washing, myocytes were also incubated with MitoTracker Deep Red (#M46753, ThemoFisher Scientific) in HBSS buffer for 15 minutes at 37°C to identify the mitochondria in the live cells. Myocytes were visualized in HBSS buffer containing calcium and magnesium. Images were generated with a laser scanning confocal microscope Leica SP5 and an inverted microscope with a 63x1.3 objective using Leica LAS AF acquisition software. The MitoSOX fluorescence intensity was measured by transferring MitoTracker-originated regions of interest (blue channel) to a MitoSOX image (red channel) using ImageJ Fiji (1.54f) and the built-in Analyze Particle function. The intensity values were normalized to *Erbb4*$^{fl/+}$ myocytes per the assay.

**Transmission electron microscopy (TEM)**

A standard procedure for TEM of the adult mouse heart was utilized by the Microscopy Core at New York University Grossman School of Medicine. Briefly, exposed hearts in the thoracic cavity in experimental mice were retrogradely perfused with 4% PFA in 0.1 M PBS, then scarified, and the hearts were dissected. LA was further dissected and stored in Eppendorf tube containing freshly prepared 2.5% glutaraldehyde, 2% paraformaldehyde in 0.1M phosphate buffer (PB), pH 7.2 at room temperature for 1 hour. Subsequently, LA tissue was further dissected to 1 mm pieces and submerged into the fresh fixative and fixed at 4 °C overnight. After washing with 0.1M PB, samples were postfixed in 1% osmium tetroxide (OsO4) with 1% potassium ferrocyanide in 0.1M PB for 1.5 hours on ice. Followed by washing steps, samples were dehydrated in a graded series of ethanol (30%, 50%, 75%, 85%, 90%, 100%:100%), infiltrated with propylene oxide and EMbed 812 (Electron Microscopy Sciences, PA) and embedded according to a standard protocol. 70 nm thin sections were cut with a Leica Ultracut UC6 ultramicrotome (Leica, Vienna, Austria) and collected on 200 mesh copper grids (Ted Pella, Inc. CA), stained with uranyl acetate and lead citrate, and subsequently examined with a JEOL1400 Flash Transmission EM (JEOL, Japan) operated at an accelerating voltage of 120kV. Digital images were recorded with a Gatan Rio16 camera (Gatan Inc. CA). The longitudinal area of mitochondria was measured using ImageJ Fiji (1.54f).

**Supplemental Figure S1. ERBB4 expression in adult human and mouse heart.**

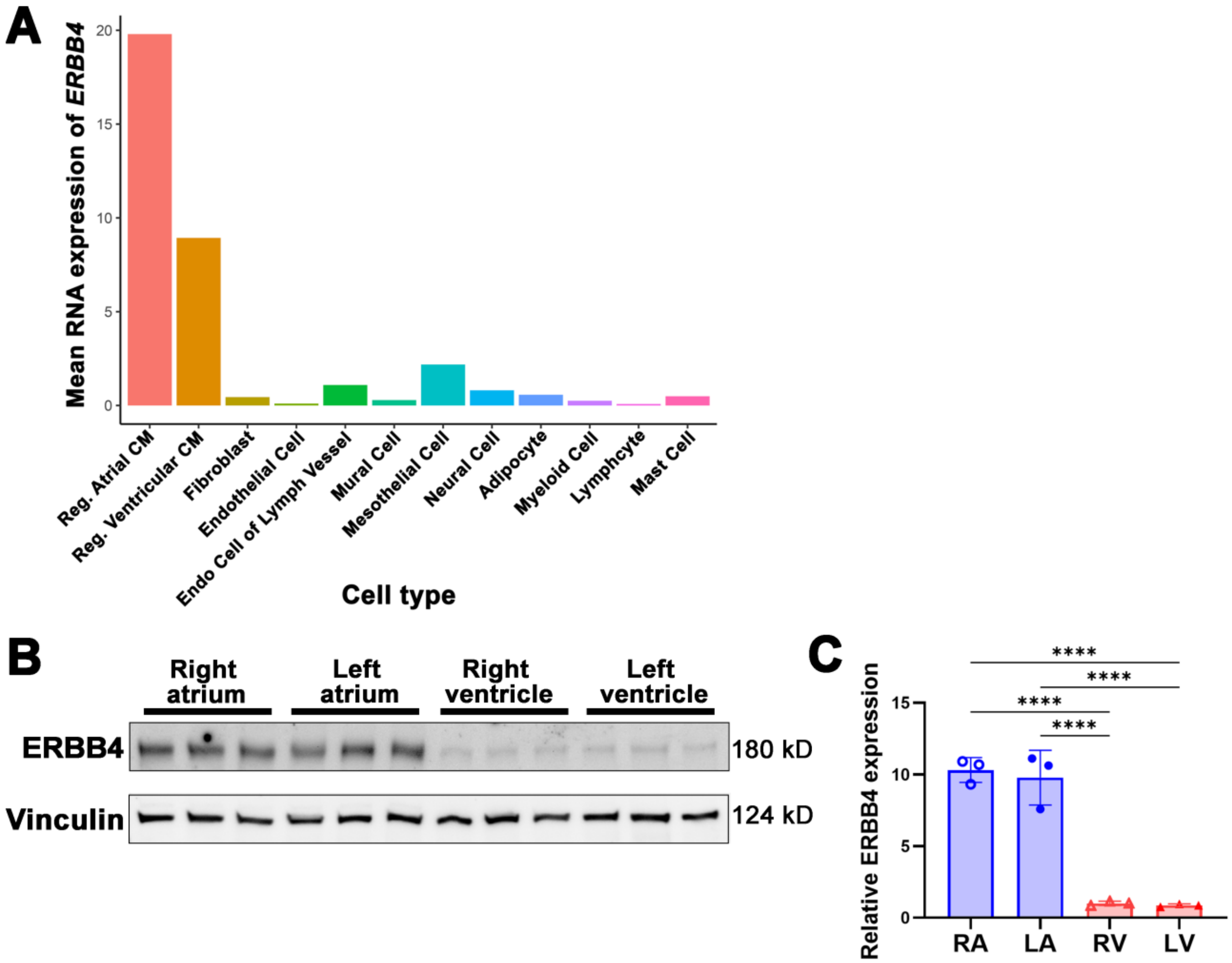


**A**, Mean *ERBB4* expression levels in individual cell types of the adult human heart. Reg. Atrial CM, regular atrial cardiac myocyte; Reg. Ventricular CM, regular ventricular cardiac myocyte; Endo Cell of Lymph Vessel, endothelial cell of lymphatic vessel. **B,** Representative immunoblot with wildtype adult mouse heart and **C**, Quantification of ERBB4 protein expression from (B) showing higher ERBB4 expression in the atria compared to the free wall of ventricles. n=3 biological replicates per tissue. Values are normalized to right ventricle (RV) and presented as mean±SD. ****$P$<0.0001 by one-way analysis of variance test followed by Tukey's post hoc test.

**Supplemental Figure S2. Cardiac axis and echocardiographic assessment in *Erbb4*$^{fl/+}$Mlc2a$^{Cre}$ and control mice.**

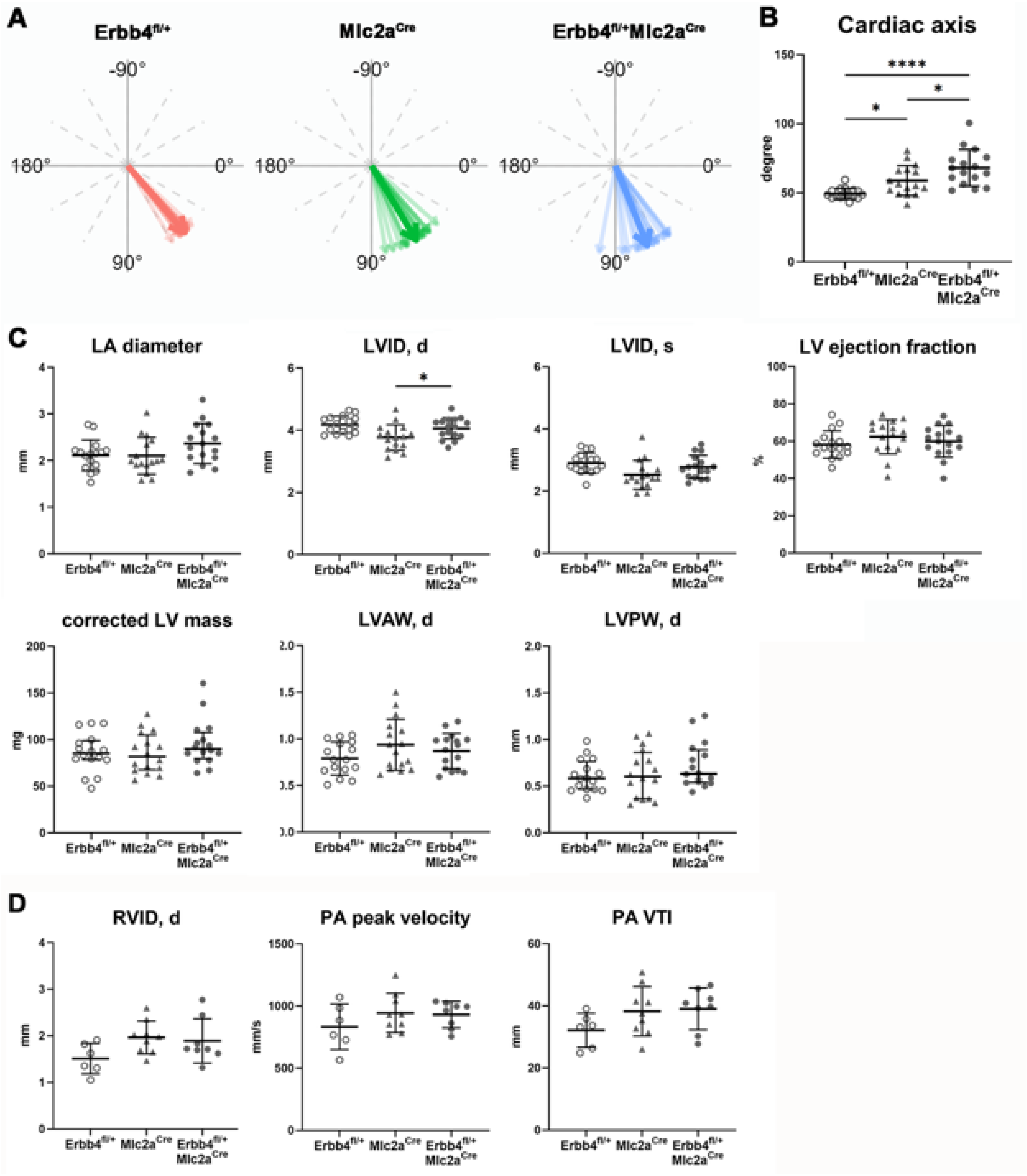


**A**, Frontal cardiac electrical axes of individual mice (light arrows) and mean group axes (dark arrows) derived from lead I and lead II. **B**, Group comparisons of cardiac axis.

Values are presented as mean±SD. n = 16 per group. *$P<0.05$, ****$P<0.0001$ by one-way analysis of variance test followed by Tukey's post hoc test. **C**, Comparisons of transthoracic echocardiography parameters of LA and left ventricle (LV). Values are presented as mean±SD; corrected LV mass and LVPW,d are presented in median with interquartile range. *$P<0.05$ by one-way ANOVA followed by Tukey's post-hoc test. Corrected LV mass and LVPW,d compared by Kruskal-Wallis test. n = 16 mice per group. LA, left atrium; LVID,d, diastolic left ventricular internal diameter; LVID,s, systolic left ventricular internal diameter; LVAW,d, diastolic left ventricular anterior wall thickness; LVPW,d, diastolic left ventricular posterior wall thickness. **D**, Comparisons of transthoracic echocardiography parameters for right ventricular morphology and function showing no significant difference between groups. Data are shown as mean±SD. n = 6−9 per group. RVID,d, diastolic right ventricular internal diameter; PA, pulmonary artery; PA VTI, pulmonary artery velocity time integral.

**Supplemental Figure S3. ERBB4 reduction leads to changes in expression of other atrial fibrillation-risk genes in mouse left atria with corresponding correlations in human left atria**

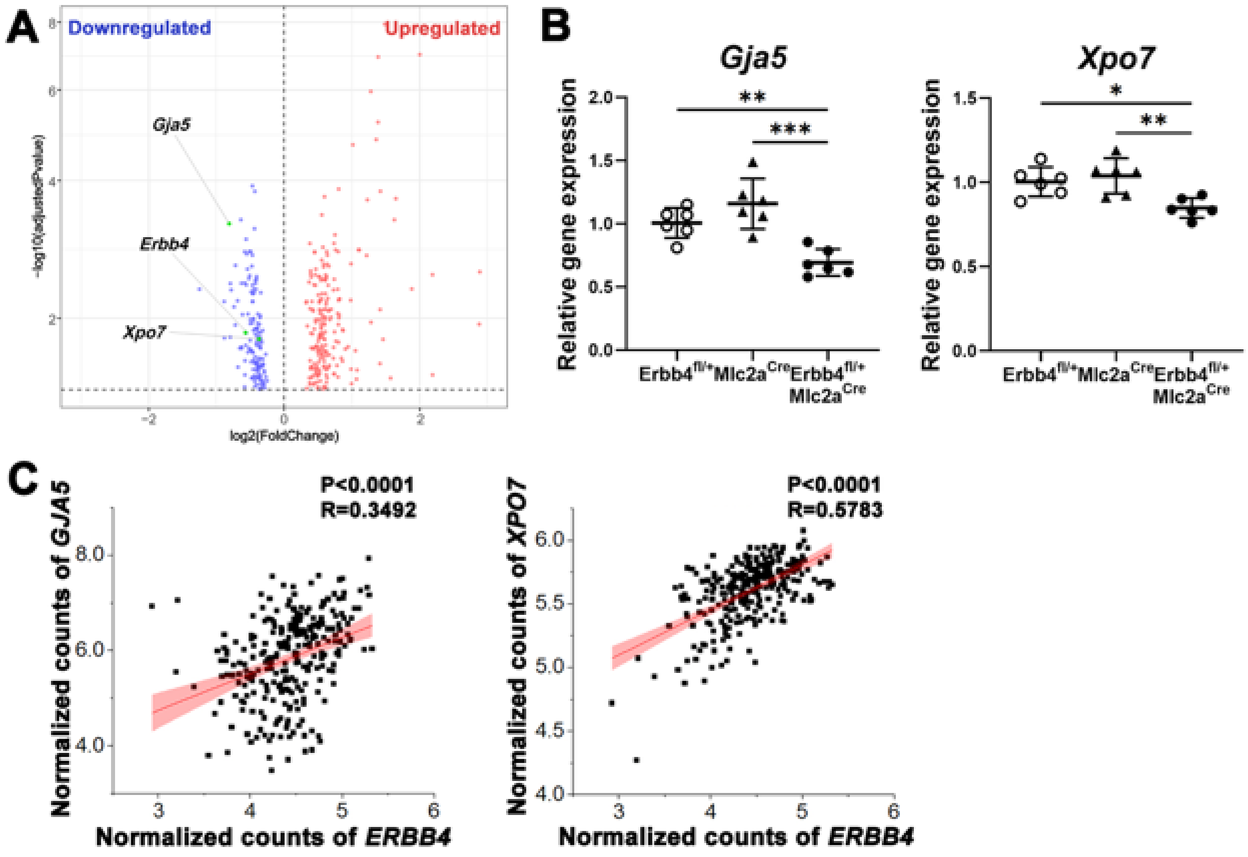


**A**, Volcano plot of 362 differentially expressed genes (adjusted *P* value <0.05) in *Erbb4*$^{fl/+}$Mlc2a$^{Cre}$ mouse left atria. Dots in red represent significantly upregulated genes and dots in blue represent downregulated genes. Atrial fibrillation (AF) risk genes by GWAS are highlighted in green. *Erbb4*, *Gja5*, and *Xpo7* are downregulated. **B**, RT-qPCR validation of selected AF-risk genes in mouse left atria. Data are shown as mean±SD. *$P$<0.05, **$P$<0.01, ***$P$<0.001 by one-way ANOVA followed by Tukey's post-hoc test. **C**, *ERBB4* expression in human LA correlates with the AF risk genes, *GJA5* and *XPO7*, by simple linear regression analysis in the Cleveland Clinic Biobank dataset.

**Supplemental Figure S4. Intracellular signaling downstream of ERBB4 in left atria.**

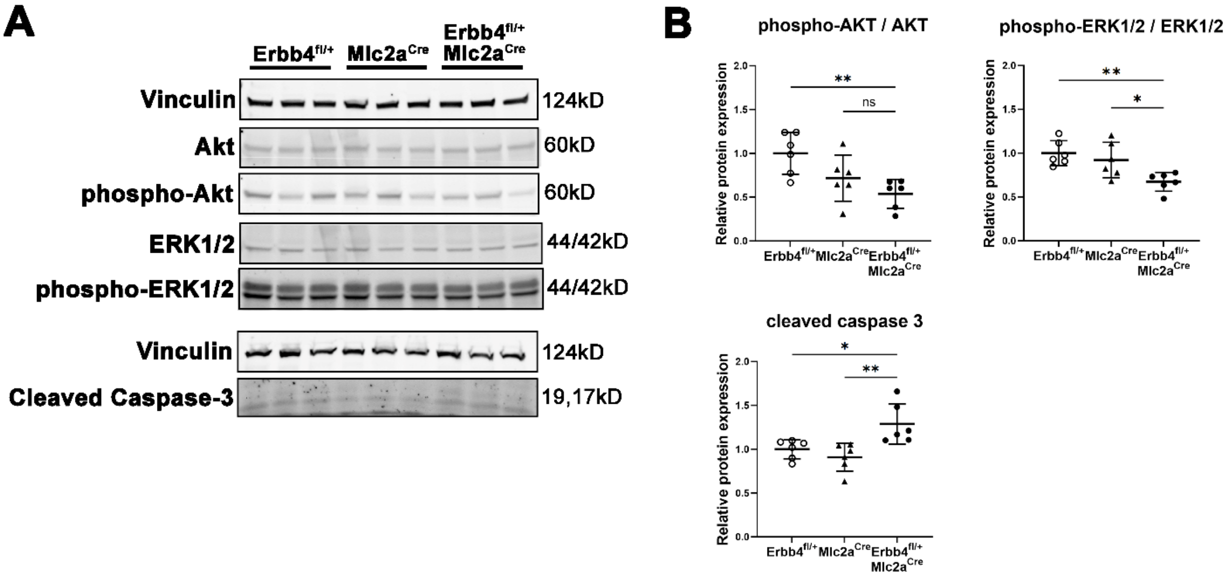


**A**, Representative immunoblot of AKT, phospho-AKT, ERK1/2, phospho-ERK1/2, and cleaved caspase-3 in left atrial lysates from *Erbb4*$^{fl/+}$Mlc2a$^{Cre}$ and control mice. Upper and lower blot panels were run on separate membranes; Vinculin (124 kDa) served as the loading control for each. **B**, Densitometric quantification of relative protein expression. Data are shown as mean±SD. n = 6 per group. *P<0.05, ***P*<0.01 by one-way analysis of variance test followed by Tukey's post hoc test.

**Supplemental Figure S5. Human left atrial *ERBB4*-correlated genes behave similarly to differentially expressed genes in the *Erbb4* haploinsufficiency mouse model**

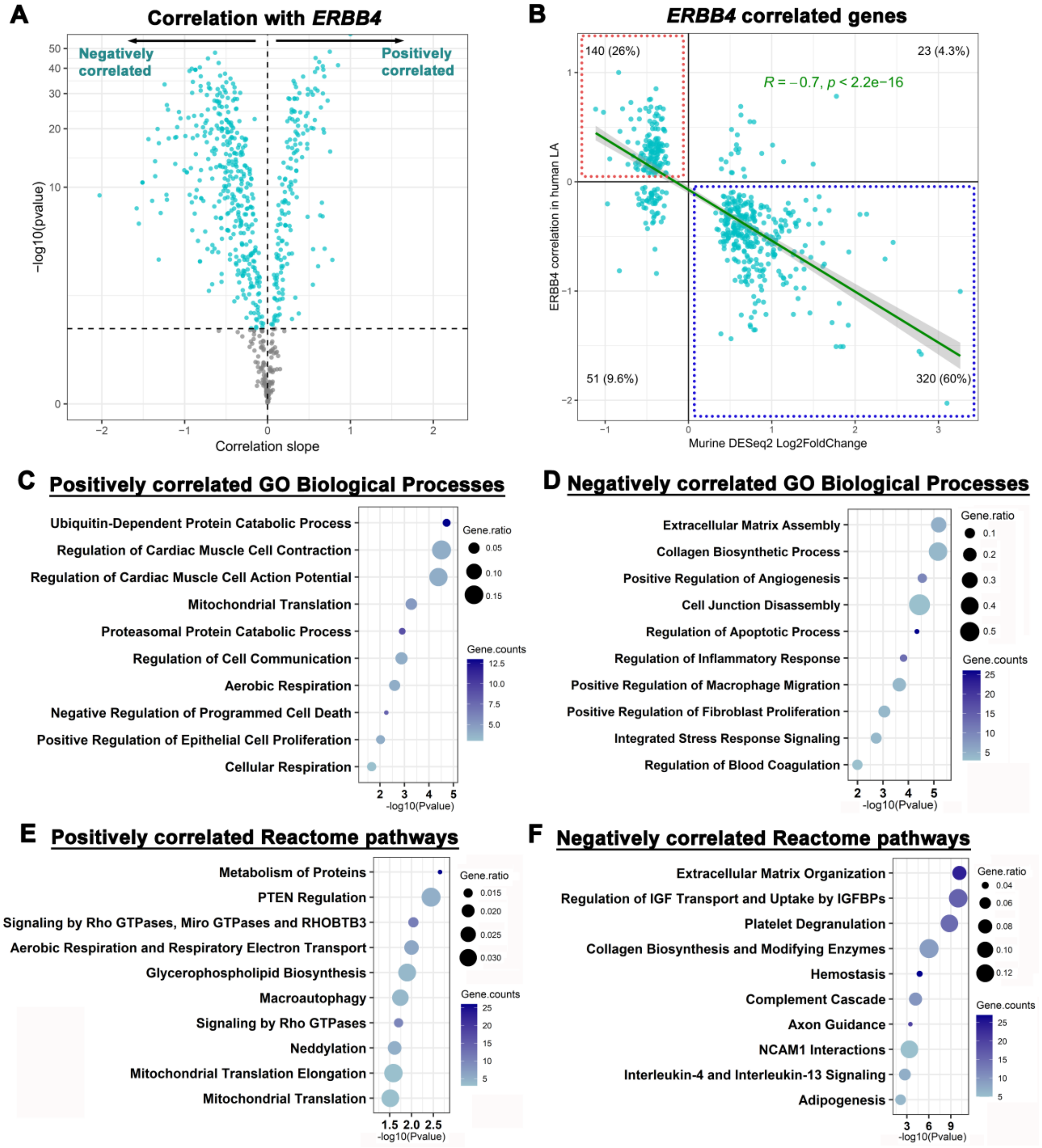


**A**, Volcano plot showing genes positively and negatively correlated with *ERBB4* expression in the human left atrial dataset. Human orthologs (n = 646) of differentially expressed genes (DEGs) in the *Erbb4* haploinsufficiency mouse model (*Erbb4*$^{fl/+}$Mlc2a$^{Cre}$) were analyzed. Candidate genes associated with *ERBB4* are highlighted in cyan (*P* value <0.05, n = 534). **B**, Scatter plot comparing the expression pattern of *ERBB4*-associated orthologs (n = 534) in human left atria (LA) with DEGs in *Erbb4*$^{fl/+}$Mlc2a$^{Cre}$ mice. The green line represents the Pearson correlation between human and mouse datasets. Genes within the dashed boxes (red: upper-left; blue: lower-right) represent genes with directionally concordant associations in human LA (n = 460). **C–F**, Dot plots of curated subsets of significant GO Biological Processes (C, D) and Reactome pathways (E, F) using the 460 concordantly correlated genes. Gene count indicates the number of input genes annotated to each GO term or Reactome pathway. Gene ratio is a proportion of input DEGs among all annotated genes in each term/pathway. IGFBPs, insulin-like growth factor binding proteins; NCAM1, neural cell adhesion molecule-1; PTEN, phosphatase and tensin homologue; RHOBTB3, Rho Related BTB Domain Containing 3.

**Supplemental Figure S6. Reactome pathway analysis in GTEx data set**

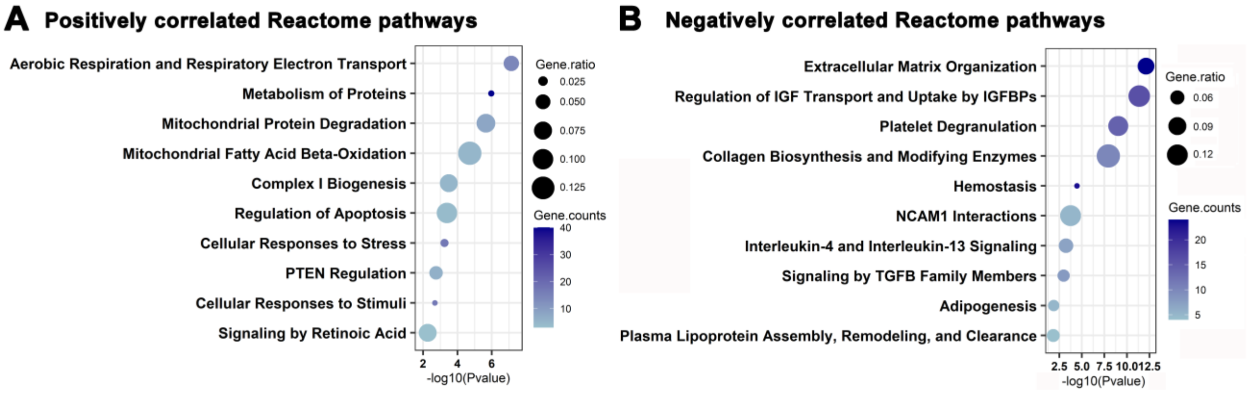


**A,B**, Dot plots of curated subsets of significant Reactome pathways identified by Enrichr using the 428 concordantly correlated genes with *ERBB4*. Gene count indicates the number of input genes annotated to each Reactome pathway. Gene ratio is a proportion of input DEGs among all annotated genes in each pathway. IGF, insulin-like growth factor; IGFBPs, insulin-like growth factor binding proteins; NCAM1, neural cell adhesion molecule-1; PTEN, phosphatase and tensin homologue; TGFB, transforming growth factor beta.

**Supplemental Figure S7. Co-staining of TUNEL and cytoskeleton marker α-actinin**

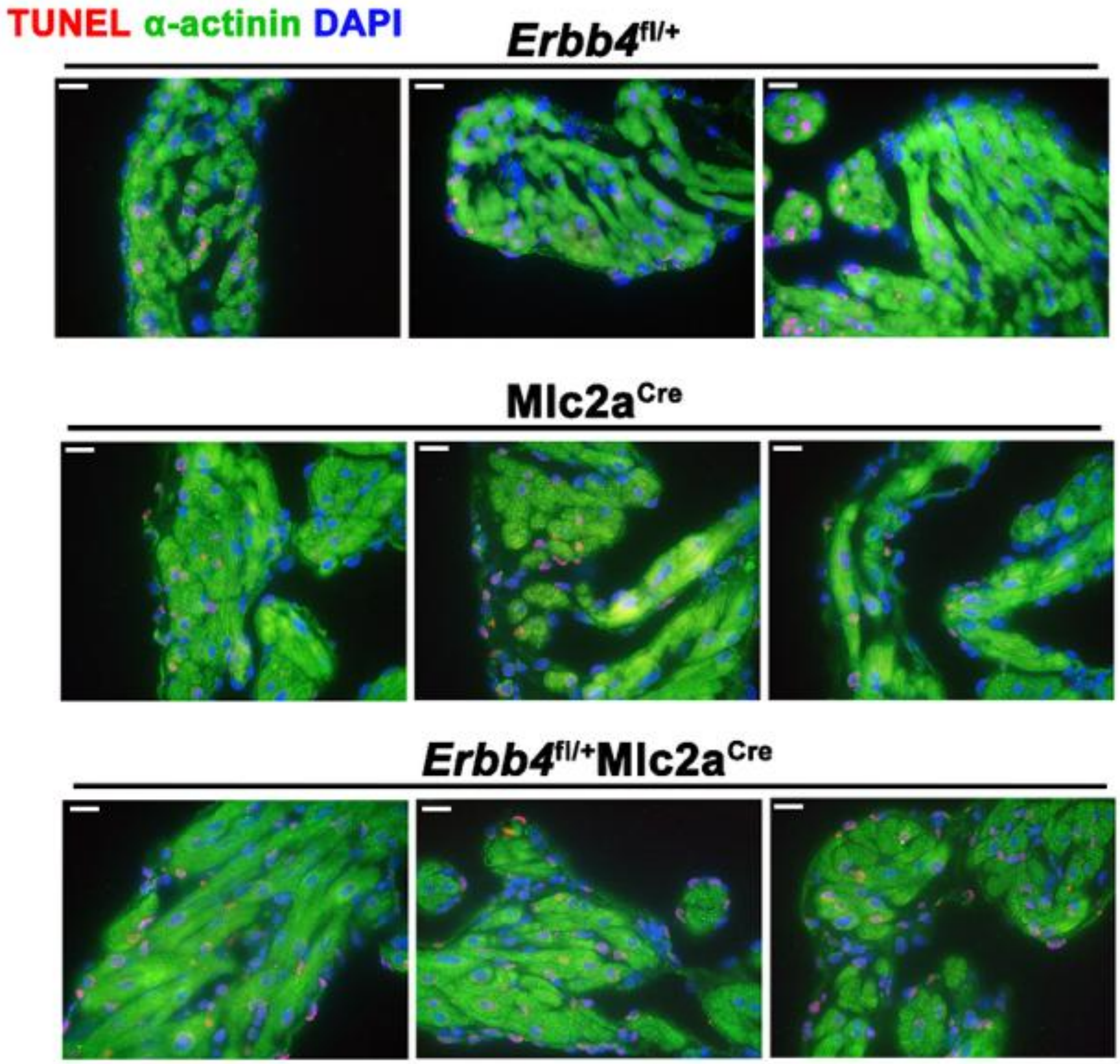


Representative images of terminal deoxynucleotidyl transferase dUTP nick end labeling (TUNEL) assay sections counterstained with cytoskeleton marker α-actinin 1 (ACTN1) and a DNA-specific dye, 4′,6-Diamidino-2-phenylindole dihydrochloride (DAPI). Scale bar = 20 μm. These images were not included in the quantitative analysis shown in Figure 6D.

**Supplemental Figure S8. Ultrastructural assessment in *Erbb4*$^{fl/+}$Mlc2a$^{Cre}$ and control mouse LA**

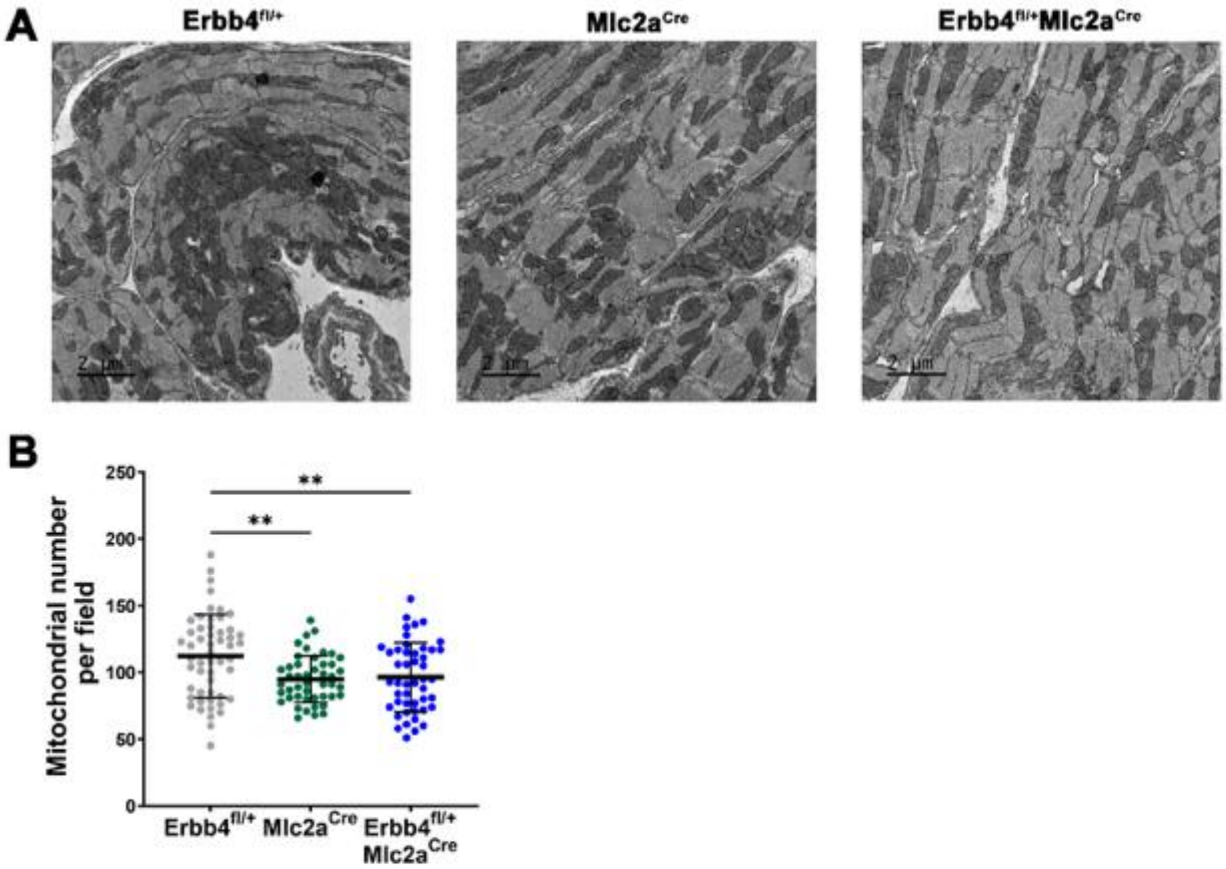


**A**. Representatives transmission electron microscopy images of left atrial sections in the mouse models. **B**. Mitochondrial number per field was analyzed with ImageJ FIJI software. Total 46–53 fields were analyzed from n = 3 animals per group. Data are shown as mean±SD. ***P*<0.01 by one-way analysis of variance test followed by Tukey's post hoc test.
